# Influence of Sex and Race/ Ethnicity on Major Adverse Cardiovascular Outcomes Following Coronary Artery Bypass Surgery in a Large Integrated Healthcare System

**DOI:** 10.1101/2025.02.18.25322505

**Authors:** Janine Y. Yang, Douglas Stram, John Doan, Alix P. Fairman, Maqdooda Merchant, Cynthia Triplett, Ahmad Y. Sheikh, Richard V. Ha, Seema K. Pursnani

## Abstract

**Background:** We aimed to evaluate the influence of sex and race/ ethnicity on major adverse cardiovascular events (MACE) following coronary artery bypass grafting (CABG) in our integrated healthcare system.

**Methods:** This was a retrospective study of Kaiser Permanente Northern California members who underwent CABG from 2008-2019, evaluating odds of MACE (MI, stroke, serial percutaneous intervention (PCI), repeat CABG, death) at 30 days, 1 year, and up to 12 years follow-up using multivariable logistic and Cox proportional regression analyses. We adjusted for demographic, clinical, socioeconomic risk factors, and surgical characteristics.

**Results:** Cohort included n=7405, mean age 65.2 yrs, 47% diabetic, 62% hypertensive, 20% with prior revascularization (PCI or CABG). There were n=6082 males and n=1323 females with 2179 (35.8%) and 639 (48.3%) MACE, respectively. MACE occurred in 40.4% of White, 38.1% of Hispanic, 32.9% of Filipino, 27.9% of South Asian, 29.1% of Other Asian/ Pacific Islander, 47.0% of Black, and 42.3% of Other Race/ Ethnicity patients (p<0.001). Older age, higher HbA1c, diabetes, end-stage renal disease, lower hemoglobin, higher creatinine, smoking, lack of cardiopulmonary bypass, and use of non-arterial graft were significant predictors of long-term MACE. Female sex was associated with an increased odds of MACE at 30 days (OR 1.62, 95% CI, 1.19-2.21) and 1-year (HR 1.24, 95% CI, 1.02-1.51). Asian race/ ethnicity was associated with lower 12-year hazard of MACE (HR Filipino 0.72; 95% CI, 0.60-0.87; South Asian 0.72, 95% CI, 0.50-1.03; Other Asian 0.71, 95% CI, 0.58-0.87).

**Conclusion:** Female, Black, and Other Race/ Ethnicity groups had the greatest incidence of MACE at up to 12 years follow-up post CABG. These differences are largely driven by increased risk factor burden in these groups, and Black and Other race/ ethnicity were not independently associated with long-term CABG risk in multivariable modeling. Further understanding of the mechanisms for these sex and race/ethnic differences is required to improve upstream preventive efforts.

## Background

Ischemic heart disease remains the leading cause of death in the United States and in the world.^1,2^ In the treatment of ischemic heart disease, coronary artery bypass graft (CABG) surgery is often preferred over percutaneous coronary intervention (PCI) for revascularization in cases with multivessel disease or left main coronary artery disease (CAD), especially in those with complex anatomy and concurrent diabetes mellitus (DM).^2,3^ Although advancements in surgical techniques and medical therapy have significantly reduced CABG mortality rates, disparities in treatment access and surgical outcomes persist.^4^

Prior studies have demonstrated that female and non-White racial/ ethnic groups have worse cardiovascular outcomes following CABG.^5–12^ Data from the National Inpatient Sample from 2002-2017 demonstrated that Black patients had the lowest odds of undergoing CABG as well as the highest adjusted mortality rate following CABG along with Hispanic patients.^13^ These disparities are only partially explained by patient risk factors, hospital quality, and socioeconomic status.^14^ Some of these differences may be explained by deviations in perioperative management. For example, Black and Hispanic patients are less likely to receive arterial grafts during CABG, which have been shown to have better long-term patency as conduits compared to venous grafts.^15–18^ In Asian populations, CABG operative mortality is similar to White populations, despite South Asians carrying an increased burden and severity of cardiovascular risk factors.^19,20^ While South Asians are more likely to have DM, hypertension (HTN), and dyslipidemia compared to other racial/ ethnic groups, South Asian ethnicity itself is not associated with adverse post CABG outcomes in multivariable modeling.^19–24^ Sex differences in post CABG outcomes also exist due to a higher burden of preoperative risk factors and variations in CAD progression.^6,25,26^ Female patients undergoing CABG are typically older with a higher prevalence of DM, HTN, peripheral arterial disease, and dyslipidemia compared to males.^27–29^ Female patients are also more likely to present with decompensated heart failure, acute myocardial infarction or cardiogenic shock preceding CABG.^27–29^ Even after adjusting for traditional risk factors, female patients undergoing CABG experience higher operative mortality and are at a greater risk of major postoperative complications such as myocardial infarction and stroke.^27,29^

Further contemporary understanding of racial/ ethnic and sex differences in baseline risk factors and outcomes in patients undergoing CABG is required to better inform clinical decision-making and reduce health disparities. In this study, we aim to evaluate the influence of race/ ethnicity and sex on short- and long-term major adverse cardiovascular events (MACE) following CABG in our diverse integrated healthcare system.

## Methods

We performed a retrospective cohort study of Kaiser Permanente Northern California (KPNC) members who underwent isolated CABG between 2008 and 2019. Patients for whom self-reported race/ ethnicity data was available were included in the study cohort. Members who lacked continuous membership at least one year following CABG for reason other than death were excluded. Demographic, clinical, and surgical data were extracted from the Society of Thoracic Surgeons (STS) Database and KPNC electronic health record system. Baseline data collected included demographics (self-reported race/ ethnicity, sex, census tract education and income level, primary language), medical risk factors (smoking, body mass index, baseline glycated hemoglobin (HbA1c), hemoglobin, creatinine, low density lipoprotein cholesterol (LDL-c) and high density lipoprotein cholesterol (HDL-c), diabetes mellitus (DM), hypertension (HTN), end stage renal disease (ESRD), prior CAD) and lastly CABG characteristics (year of surgery, STS risk score, elective versus urgent surgical status, use of cardiopulmonary bypass, presence and number of arterial grafts). Temporal data collected during follow-up periods included statin use, HbA1c and LDL-c levels.

MACE was defined as myocardial infarction (MI), surgical or percutaneous coronary revascularization, stroke, and all-cause mortality. In patients who had more than one of these events during the study follow-up period, only their first MACE event was considered. MACE was evaluated at 30 days, 1 year, and up to 12 years follow-up. We performed bivariate analyses comparing baseline characteristics by sex, testing for statistical significance using chi-square tests for categorical variables and Wilcoxon rank-sum tests for continuous variables. We also compared baseline characteristics by race/ ethnicity, using chi-square tests for categorical variables and Kruskal-Wallis tests for continuous variables. Multivariable regression analysis was used to identify the odds of MACE at 30 days, and Cox proportional regression analyses were used to evaluate the hazard of MACE at up to 1- and 12 years follow-up. Multivariable analysis results were adjusted for baseline clinical and demographic characteristics. Outcomes were extracted based on documented International Classification of Diseases (ICD) and Current Procedural Terminology (CPT) codes. Kaplan-Meier curve estimates for MACE-free survival were generated stratifying sex and race/ ethnicity, and log-rank tests were used to assess differences in survival between groups. Statistical analyses were performed in SAS 9.4 for Windows. P-values were considered statistically significant at <0.05.

The study was approved by the Kaiser Permanente Northern California Institutional Review Board, and informed consent was not required nor obtained for this data-only study.

## Results

The study cohort included n=7450 patients who underwent CABG between 2008 and 2019. Surgical volume varied during the study period, with an overall increase from 2008-2019 (Figure 1). A majority of patients were male (n=6082, 82%) versus female, (n=1323, 18%). Patients self-identified as White (n=3782, 51%), Hispanic (n=1045, 14%), Filipino (n=827, 11%), Other Asian/ Pacific Islander (n=686, 10%), Black (n=396, 5%), and South Asian (n=258, 3%) (Figure 2A), with an increased proportion of non-White racial/ ethnic groups over the study period (Figure 2B).

**Figure 1.**
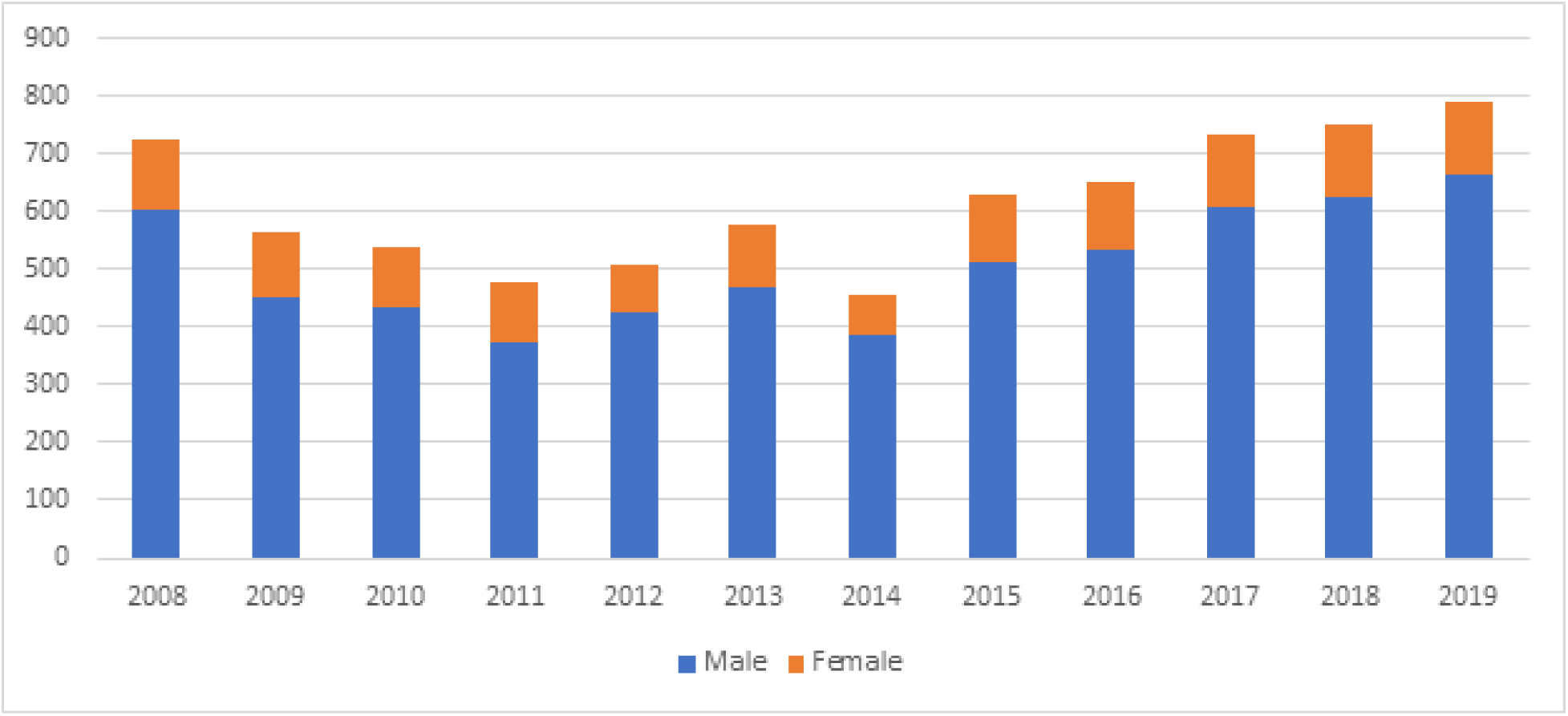
Annual number of CABG surgeries between 2008-2019 at Kaiser Permanente, Northern California. Yearly number of CABG surgeries within Kaiser Permanente, Northern California cohort are shown stratified by sex.

**Figure 2A.**
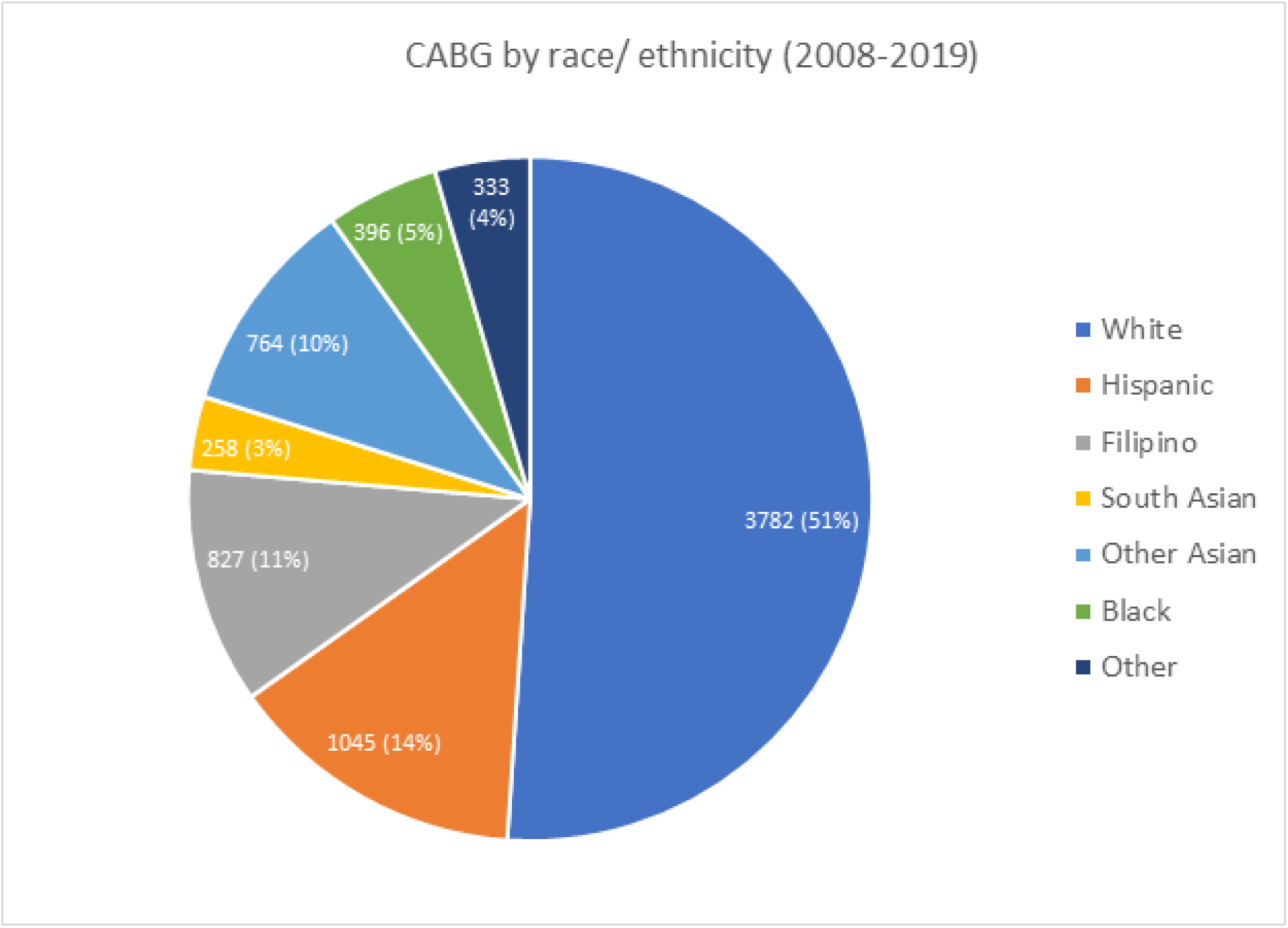
Number of CABG surgeries (2008-2019) at Kaiser Permanente, Northern California. Total number of CABG surgeries are shown, with frequency by race/ ethnicity. Other includes Native American, Multi-ethnic, and Missing race/ ethnicity groups.

**Figure 2B.**
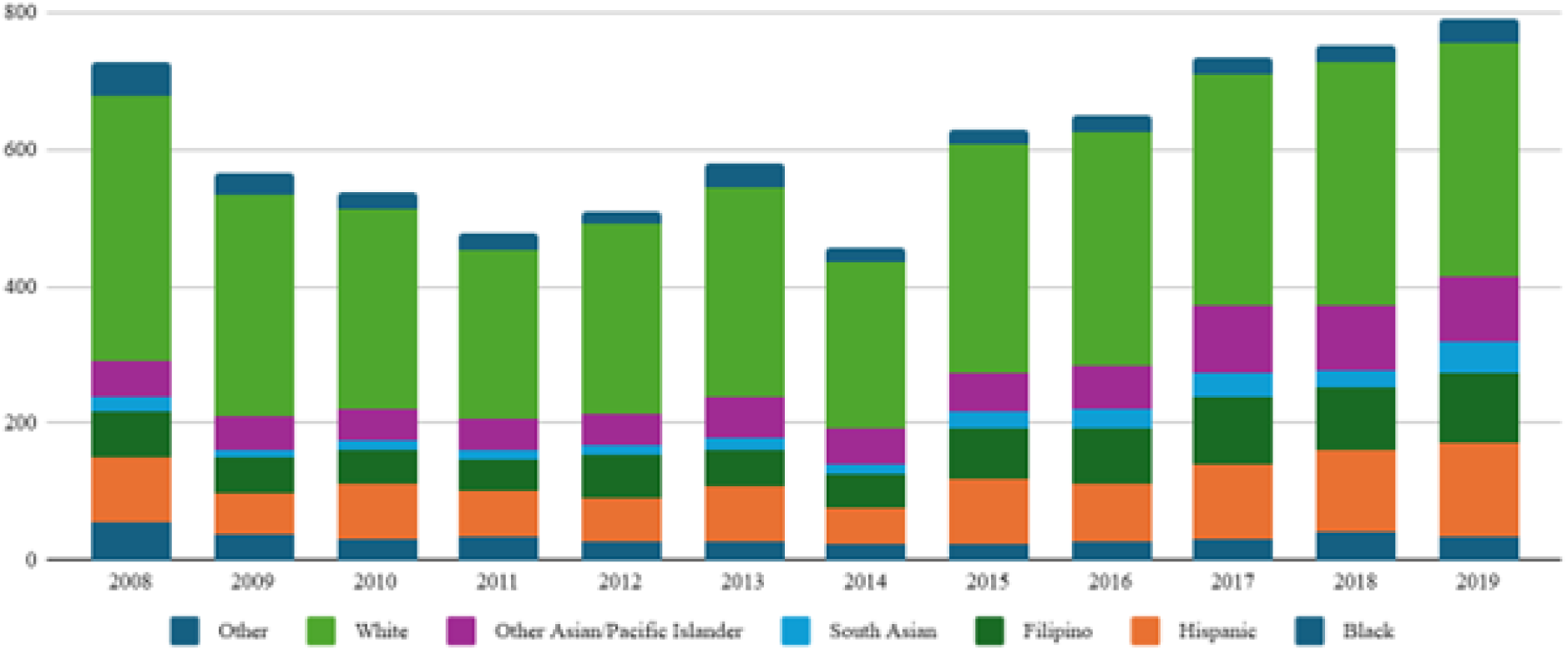
Annual number of CABG surgeries between 2008-2019 at Kaiser Permanente, Northern California, stratified by race/ ethnicity. Yearly number of CABG surgeries within Kaiser Permanente, Northern California cohort are shown stratified by race/ ethnicity. Other includes Native American, Multi-ethnic, and Missing race/ ethnicity groups.

Females, compared to males, were older at time of CABG (66.9 years versus 64.9 years, p<0.001) with higher prevalence of prior MI (58% versus 51%), DM (57% versus 45%), HTN (69% versus 61%), and ESRD (12% versus 7%), as well as higher creatinine, total cholesterol, and HbA1c values (p<0.001) (Table 1). Females had higher HDL-c levels (47.5 versus 41.4, p<0.001) and were more likely to be never smokers (57% versus 43%, p<0.0001). Females had a higher preoperative STS risk score and were more likely to undergo urgent CABG (p<0.001). Despite variation in the number of bypass grafts used, use of arterial grafts was clinically similar (male 98%, female 97%). A large majority of both males (93%) and females (92%) considered English as the primary language. Female patients lived in census tracts with a lower proportion of college graduates (42% versus 46%, p<0.001) and lower median household income ($75,565 versus $82,803, p<0.001).

**Table 1.**
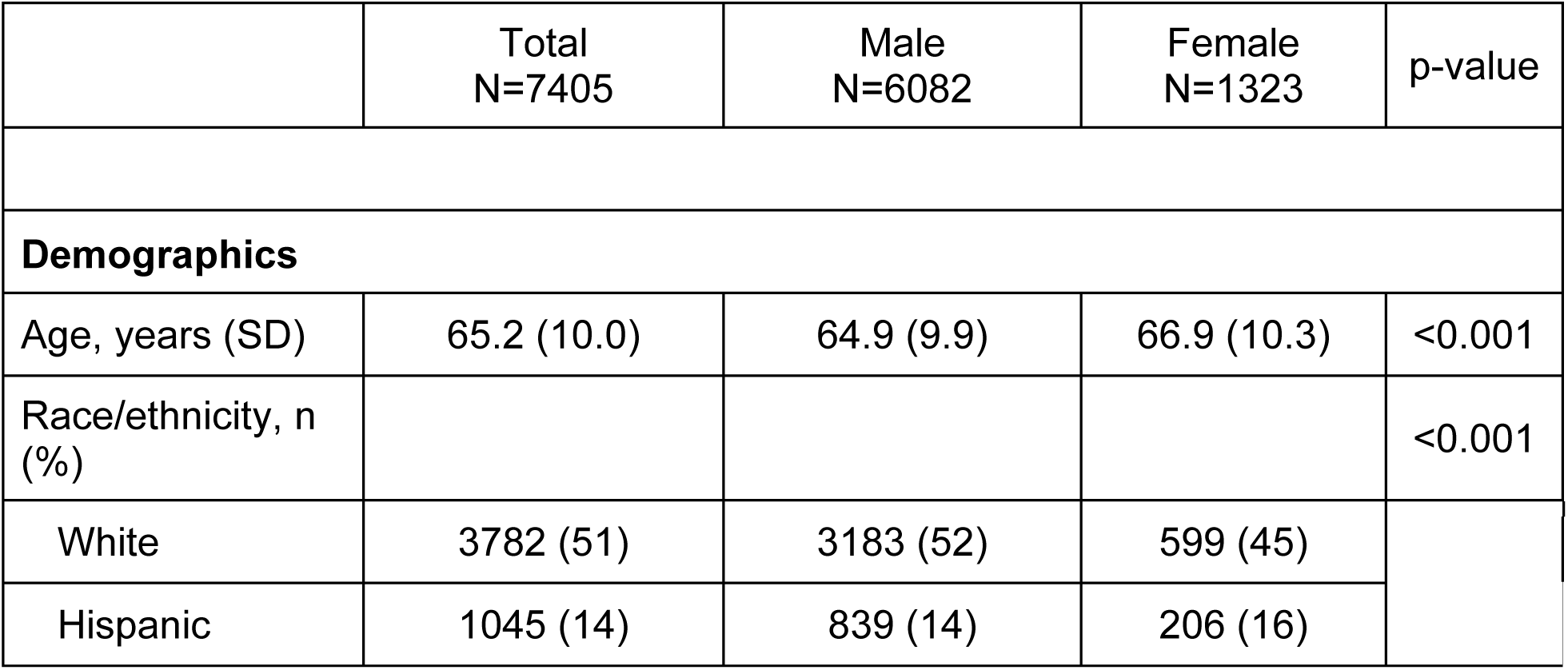
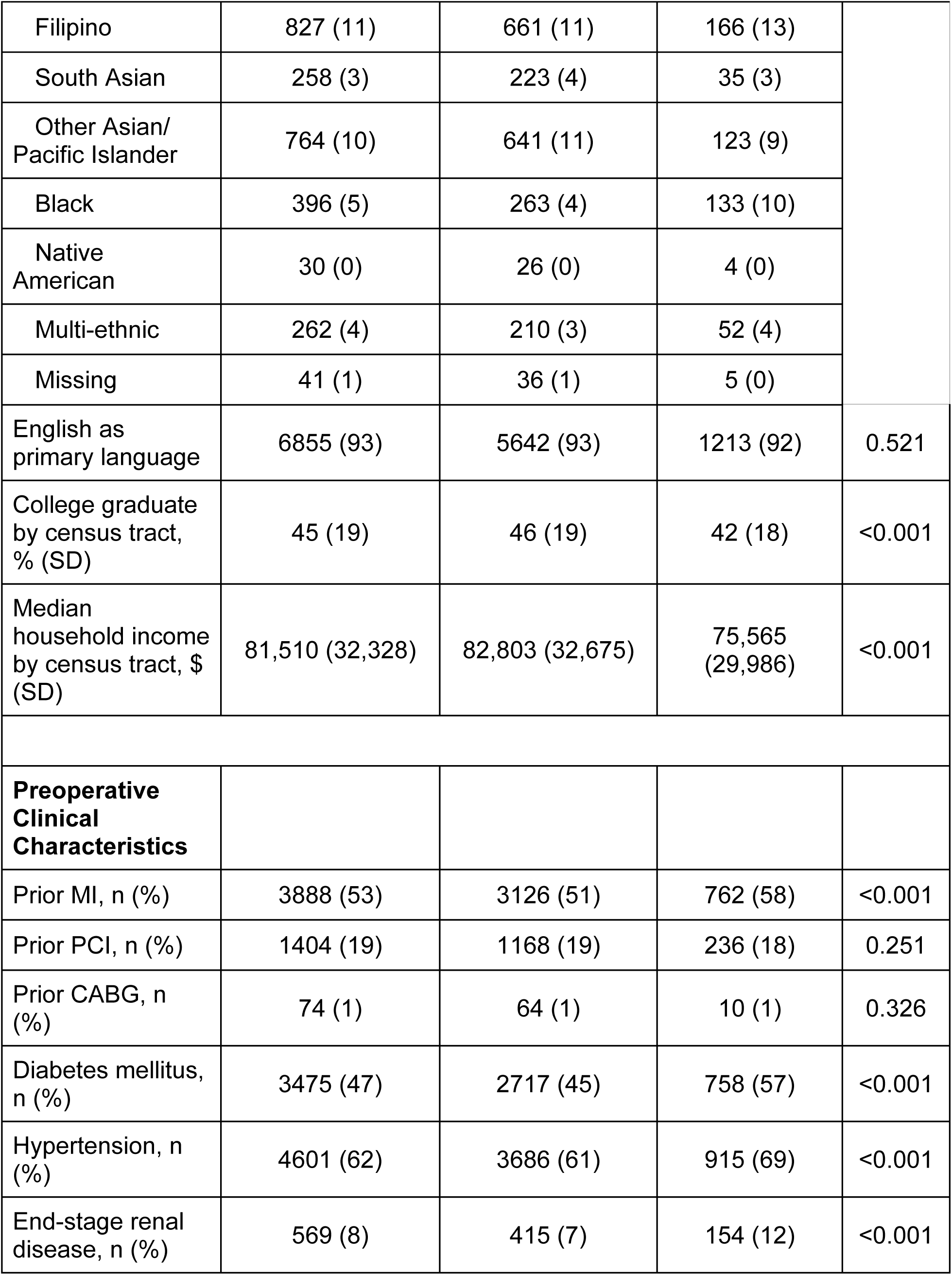
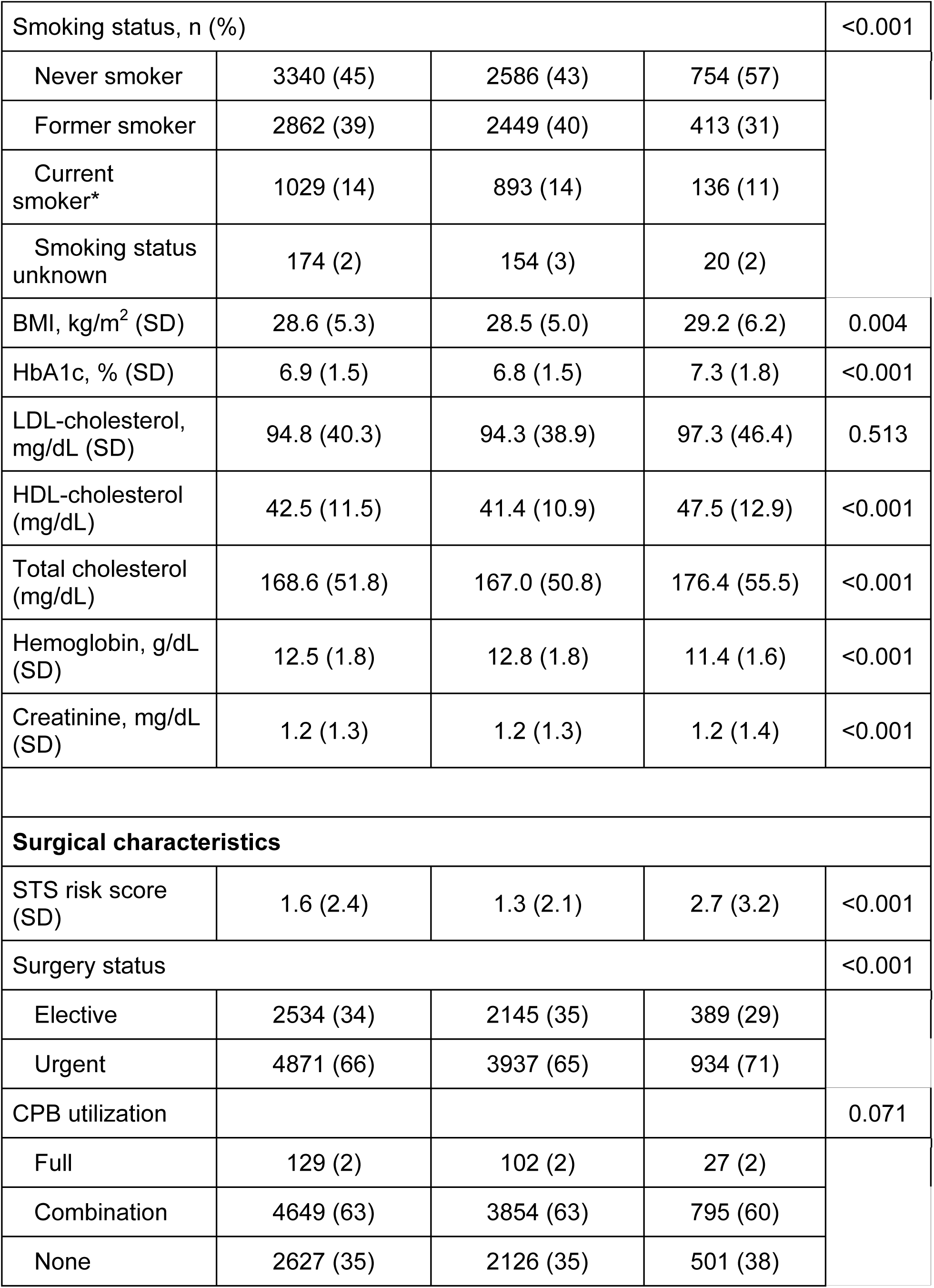
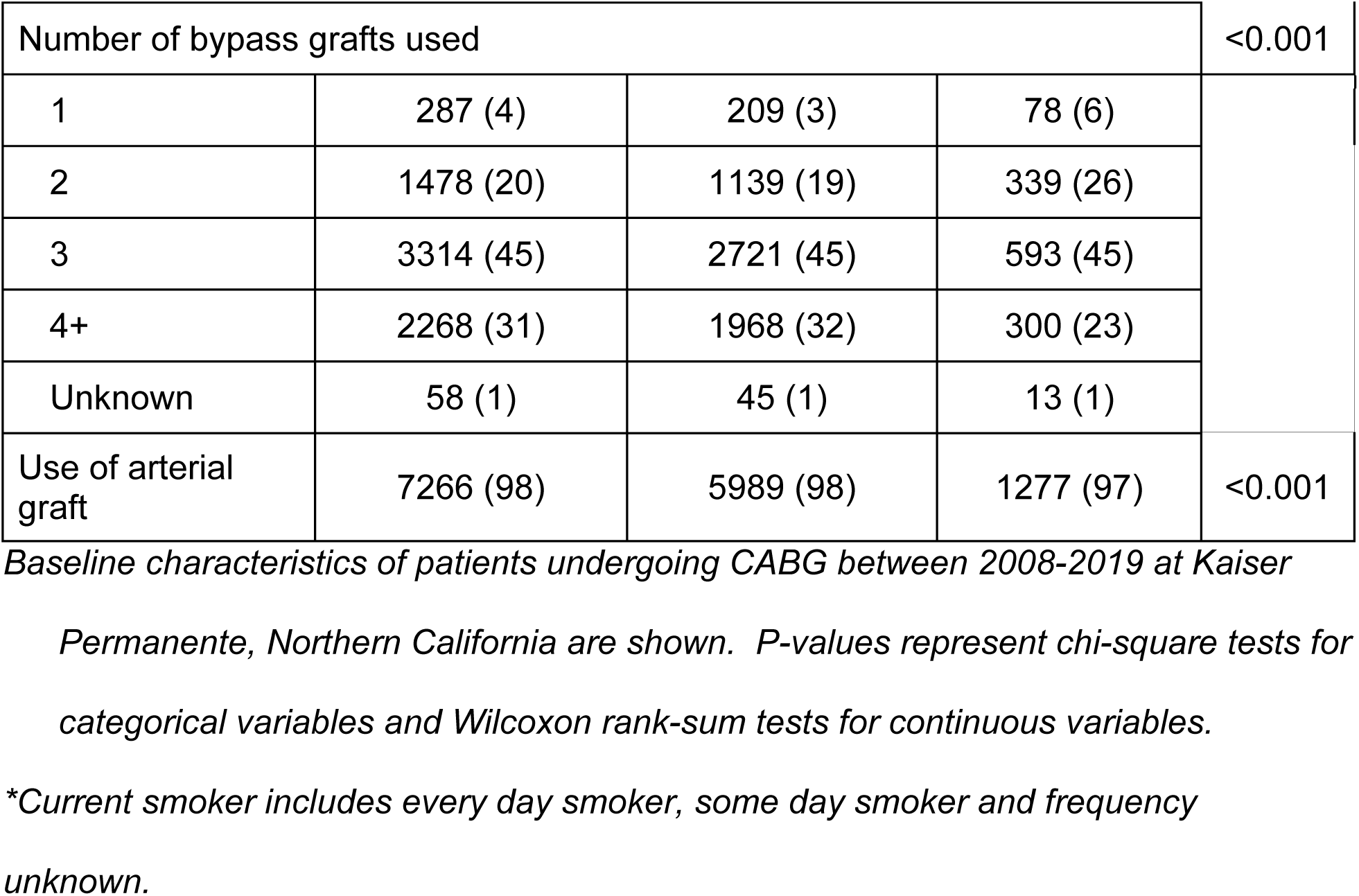
Baseline characteristics by sex.

|  | Total<br>N=7405 | Male<br>N=6082 | Female<br>N=1323 | p-value |
| --- | --- | --- | --- | --- |
| **Demographics** |  |  |  |  |
| Age, years (SD) | 65.2 (10.0) | 64.9 (9.9) | 66.9 (10.3) | <0.001 |
| Race/ethnicity, n (%) |  |  |  | <0.001 |
| White | 3782 (51) | 3183 (52) | 599 (45) |  |
| Hispanic | 1045 (14) | 839 (14) | 206 (16) |  |
| Filipino | 827 (11) | 661 (11) | 166 (13) |  |
| South Asian | 258 (3) | 223 (4) | 35 (3) |  |
| Other Asian/<br>Pacific Islander | 764 (10) | 641 (11) | 123 (9) |  |
| Black | 396 (5) | 263 (4) | 133 (10) |  |
| Native<br>American | 30 (0) | 26 (0) | 4 (0) |  |
| Multi-ethnic | 262 (4) | 210 (3) | 52 (4) |  |
| Missing | 41 (1) | 36 (1) | 5 (0) |  |
| English as<br>primary language | 6855 (93) | 5642 (93) | 1213 (92) | 0.521 |
| College graduate<br>by census tract,<br>% (SD) | 45 (19) | 46 (19) | 42 (18) | <0.001 |
| Median<br>household income<br>by census tract, \$<br>(SD) | 81,510 (32,328) | 82,803 (32,675) | 75,565<br>(29,986) | <0.001 |
| **Preoperative**<br>**Clinical**<br>**Characteristics** |  |  |  |  |
| Prior MI, n (%) | 3888 (53) | 3126 (51) | 762 (58) | <0.001 |
| Prior PCI, n (%) | 1404 (19) | 1168 (19) | 236 (18) | 0.251 |
| Prior CABG, n<br>(%) | 74 (1) | 64 (1) | 10 (1) | 0.326 |
| Diabetes mellitus,<br>n (%) | 3475 (47) | 2717 (45) | 758 (57) | <0.001 |
| Hypertension, n<br>(%) | 4601 (62) | 3686 (61) | 915 (69) | <0.001 |
| End-stage renal<br>disease, n (%) | 569 (8) | 415 (7) | 154 (12) | <0.001 |
| Smoking status, n (%) |  |  |  | <0.001 |
| Never smoker | 3340 (45) | 2586 (43) | 754 (57) |  |
| Former smoker | 2862 (39) | 2449 (40) | 413 (31) |  |
| Current smoker* | 1029 (14) | 893 (14) | 136 (11) |  |
| Smoking status unknown | 174 (2) | 154 (3) | 20 (2) |  |
| BMI, kg/m<sup>2</sup> (SD) | 28.6 (5.3) | 28.5 (5.0) | 29.2 (6.2) | 0.004 |
| HbA1c, % (SD) | 6.9 (1.5) | 6.8 (1.5) | 7.3 (1.8) | <0.001 |
| LDL-cholesterol, mg/dL (SD) | 94.8 (40.3) | 94.3 (38.9) | 97.3 (46.4) | 0.513 |
| HDL-cholesterol (mg/dL) | 42.5 (11.5) | 41.4 (10.9) | 47.5 (12.9) | <0.001 |
| Total cholesterol (mg/dL) | 168.6 (51.8) | 167.0 (50.8) | 176.4 (55.5) | <0.001 |
| Hemoglobin, g/dL (SD) | 12.5 (1.8) | 12.8 (1.8) | 11.4 (1.6) | <0.001 |
| Creatinine, mg/dL (SD) | 1.2 (1.3) | 1.2 (1.3) | 1.2 (1.4) | <0.001 |
| **Surgical characteristics** |  |  |  |  |
| STS risk score (SD) | 1.6 (2.4) | 1.3 (2.1) | 2.7 (3.2) | <0.001 |
| Surgery status |  |  |  | <0.001 |
| Elective | 2534 (34) | 2145 (35) | 389 (29) |  |
| Urgent | 4871 (66) | 3937 (65) | 934 (71) |  |
| CPB utilization |  |  |  | 0.071 |
| Full | 129 (2) | 102 (2) | 27 (2) |  |
| Combination | 4649 (63) | 3854 (63) | 795 (60) |  |
| None | 2627 (35) | 2126 (35) | 501 (38) |  |

| Number of bypass grafts used |  |  |  | <0.001 |
| --- | --- | --- | --- | --- |
| 1 | 287 (4) | 209 (3) | 78 (6) |  |
| 2 | 1478 (20) | 1139 (19) | 339 (26) |  |
| 3 | 3314 (45) | 2721 (45) | 593 (45) |  |
| 4+ | 2268 (31) | 1968 (32) | 300 (23) |  |
| Unknown | 58 (1) | 45 (1) | 13 (1) |  |
| Use of arterial graft | 7266 (98) | 5989 (98) | 1277 (97) | <0.001 |

*Baseline characteristics of patients undergoing CABG between 2008-2019 at Kaiser*
*Permanente, Northern California are shown. P-values represent chi-square tests for categorical variables and Wilcoxon rank-sum tests for continuous variables.*
*\*Current smoker includes every day smoker, some day smoker and frequency unknown.*

South Asian patients were youngest at the time of CABG (61.2 years) and White patients were oldest (67.0 years) (Table 2). South Asians were most likely to have had prior PCI (26% versus 19% in total cohort) and most likely to be never smokers (71% versus 45% in total cohort). Black individuals had the highest BMI at baseline (30.1 versus 28.6 in total cohort) and highest LDL-c levels (99.4 versus 94.8 in total cohort), as well as the lowest census tract median household income ($66,225 versus $81,510 in total cohort). Filipinos had the highest baseline creatinine levels (1.7 versus 1.2 in total cohort), highest prevalence of ESRD (17% versus 8% in total cohort), HTN (70% versus 62% in total cohort), and DM (60% versus 47% in total cohort). Hispanic patients had the highest baseline HbA1c (7.4% versus 6.9% in total cohort), lowest HDL-c levels (40.6 versus 42.5 in total cohort), the lowest proportion of patients who considered English as a primary language (74% versus 93% in total cohort), and the lowest census tract proportion of college graduates (37% versus 45% in total cohort). Black and Hispanic patients had the highest STS risk scores and were most likely to undergo urgent, as compared to elective, CABG surgery (69% in both groups, compared to 66% in total cohort).

**Table 2.**
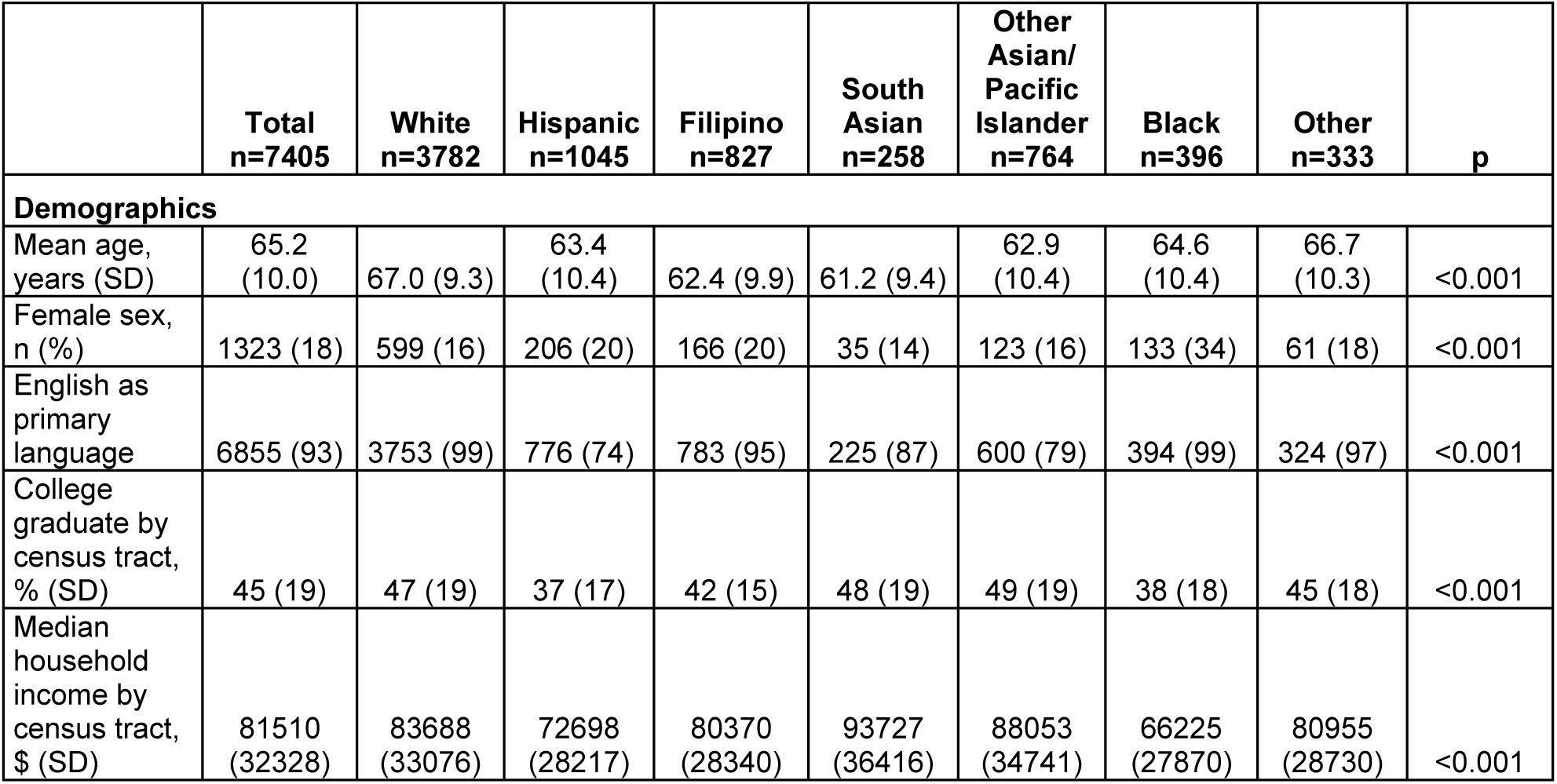
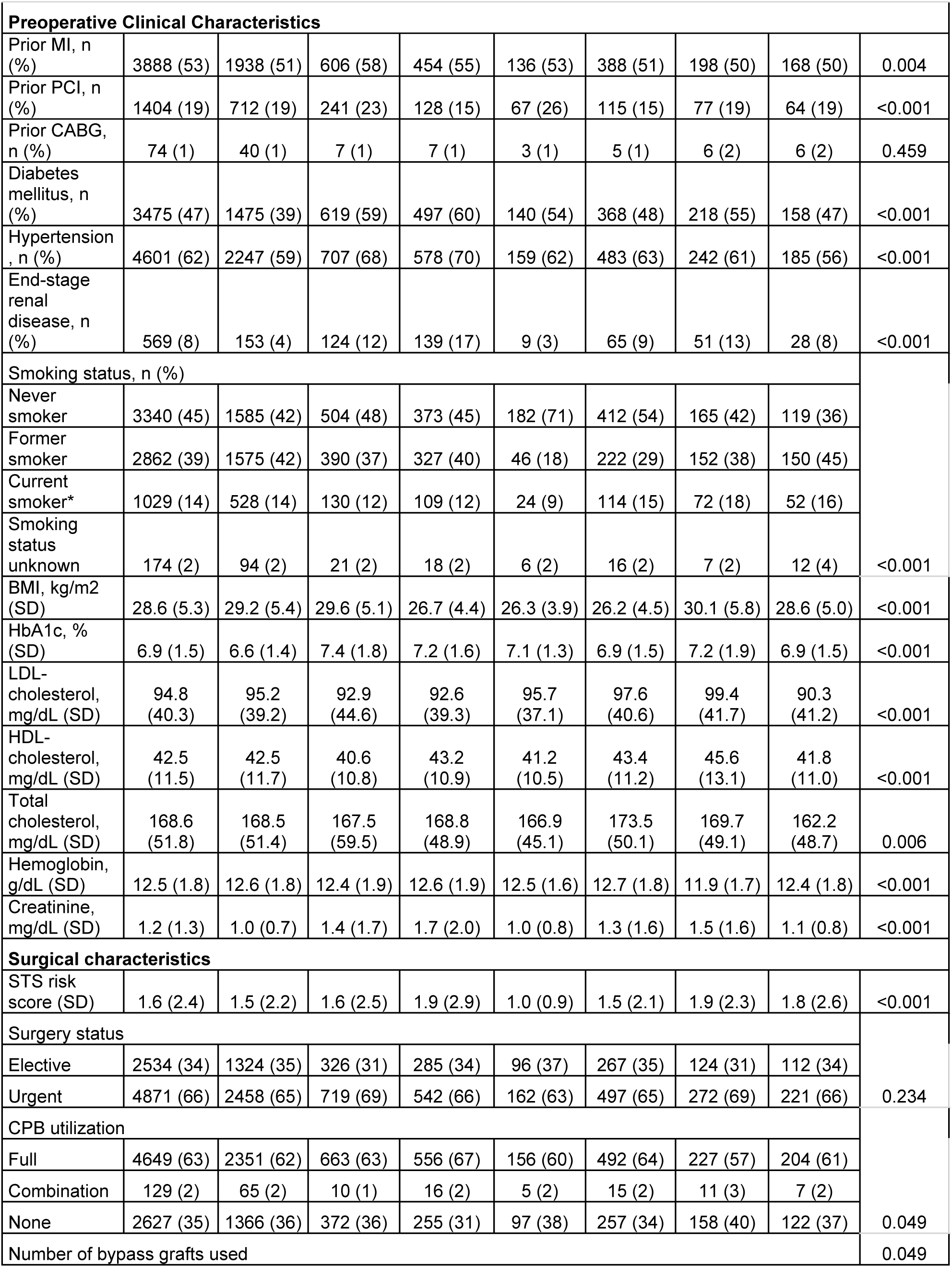
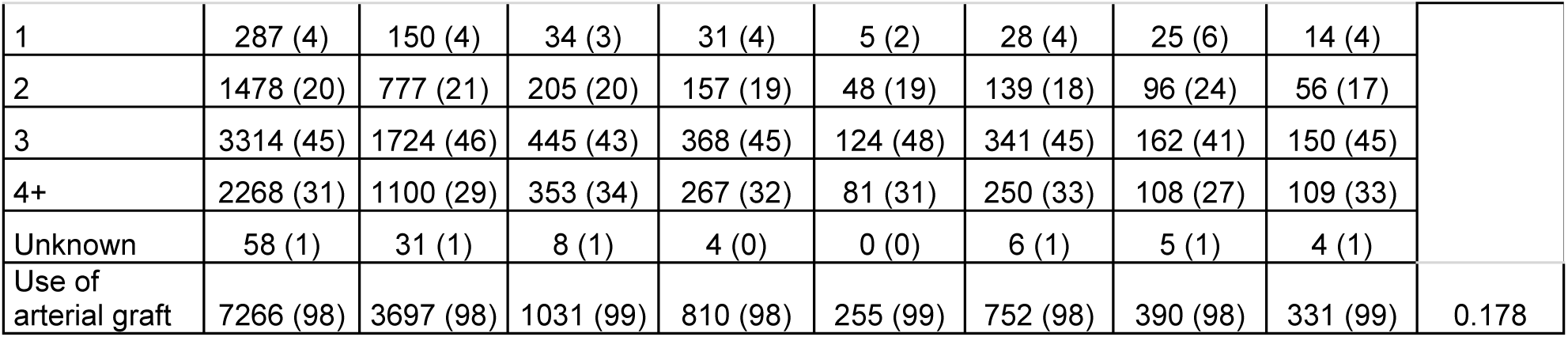
Baseline characteristics by race/ ethnicity.

In bivariate analysis, there was a strong association between MACE during 12 years of follow-up and sex (p <0.001) and race/ ethnicity (p <0.001). While no statistically significant differences were found between type of MACE event and sex, there was a statistically significant difference in type of MACE events across racial/ ethnic groups (p=0.01) (Table 3). Occurrence of any MACE was highest in Black individuals (n=186, 47%) and Other (inclusive of Native American, Multi-ethnic, Missing) race/ ethnicity groups (n=141, 42.3%), and lowest in South Asians (n=72, 27.9%) and Other Asian/ Pacific Islander (n=222, 29.1%). Black patients had the highest incidence of long-term postoperative MI (n=16, 4%) and stroke (n=92, 23.2%), and South Asians had the highest incidence of repeat revascularization (12.4% with subsequent PCI).

**Table 3.**
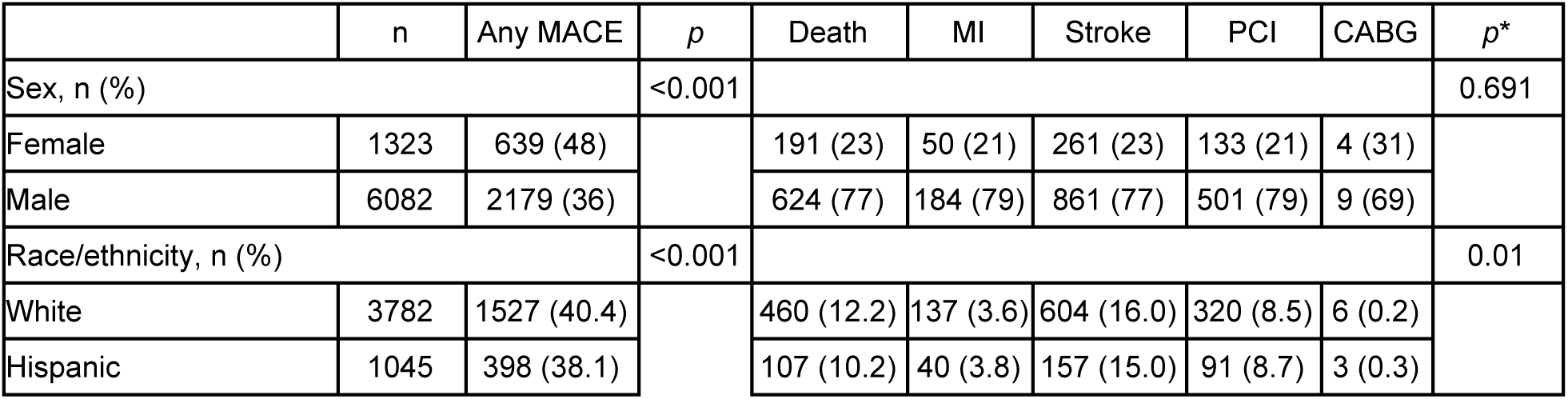
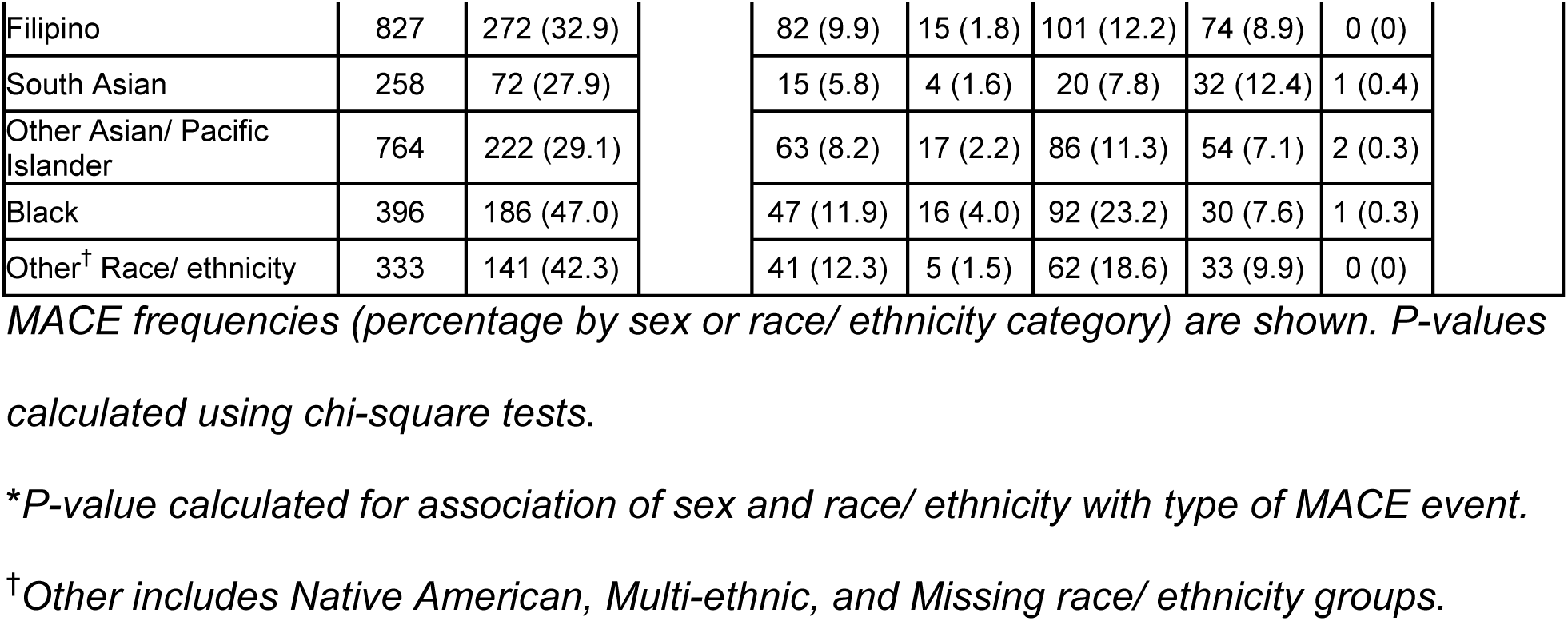
Major adverse cardiovascular events during 12 year follow up post CABG.

|  | n | Any MACE | *p* | Death | MI | Stroke | PCI | CABG | *p* * |
| --- | --- | --- | --- | --- | --- | --- | --- | --- | --- |
| Sex, n (%) |  |  | <0.001 |  |  |  |  |  | 0.691 |
| Female | 1323 | 639 (48) |  | 191 (23) | 50 (21) | 261 (23) | 133 (21) | 4 (31) |  |
| Male | 6082 | 2179 (36) |  | 624 (77) | 184 (79) | 861 (77) | 501 (79) | 9 (69) |  |
| Race/ethnicity, n (%) |  |  | <0.001 |  |  |  |  |  | 0.01 |
| White | 3782 | 1527 (40.4) |  | 460 (12.2) | 137 (3.6) | 604 (16.0) | 320 (8.5) | 6 (0.2) |  |
| Hispanic | 1045 | 398 (38.1) |  | 107 (10.2) | 40 (3.8) | 157 (15.0) | 91 (8.7) | 3 (0.3) |  |

|  |  |  |  |  |  |  |  |  |
| --- | --- | --- | --- | --- | --- | --- | --- | --- |
| Filipino | 827 | 272 (32.9) |  | 82 (9.9) | 15 (1.8) | 101 (12.2) | 74 (8.9) | 0 (0) |
| South Asian | 258 | 72 (27.9) |  | 15 (5.8) | 4 (1.6) | 20 (7.8) | 32 (12.4) | 1 (0.4) |
| Other Asian/ Pacific Islander | 764 | 222 (29.1) |  | 63 (8.2) | 17 (2.2) | 86 (11.3) | 54 (7.1) | 2 (0.3) |
| Black | 396 | 186 (47.0) |  | 47 (11.9) | 16 (4.0) | 92 (23.2) | 30 (7.6) | 1 (0.3) |
| Other<sup>†</sup> Race/ ethnicity | 333 | 141 (42.3) |  | 41 (12.3) | 5 (1.5) | 62 (18.6) | 33 (9.9) | 0 (0) |

*MACE frequencies (percentage by sex or race/ ethnicity category) are shown. P-values calculated using chi-square tests.*
*\*P-value calculated for association of sex and race/ ethnicity with type of MACE event.*
*<sup>†</sup>Other includes Native American, Multi-ethnic, and Missing race/ ethnicity groups.*

In multivariable regression analysis, variables associated with increased incidence of short-term MACE at 30-days follow-up included female sex (OR 1.62 [95% CI 1.19-2.21]), age (OR 1.03 [95% CI 1.091-1.05] per year), increased creatinine (OR 1.16 [95% CI 1.05-1.28]) and increased HbA1c (OR 1.14 [95% CI 1.04-1.25]) (Table 4). These associations with higher MACE, except for increase in HbA1c, were also preserved at 1-year follow-up. Additional variables associated with higher MACE at 1-year follow-up included DM (HR 1.28 [95% CI 1.03-1.57]), and current smoking status (HR 1.26 [95% CI 1.07-1.50]). Variables associated with decreased incidence of MACE at 30-days included Black race/ ethnicity (OR 0.44 [95% CI 0.22-0.89]) and increased hemoglobin (OR 0.85 [95% CI 0.78-0.92]), the latter which remained significantly associated with decreased MACE at 1-year follow-up (HR 0.88 [95% CI 0.84-0.93]). Surgical characteristics associated with lower MACE at 1-year follow-up included the use of arterial grafts (HR 0.59 [95% CI 0.37-0.95]).

**Table 4.**
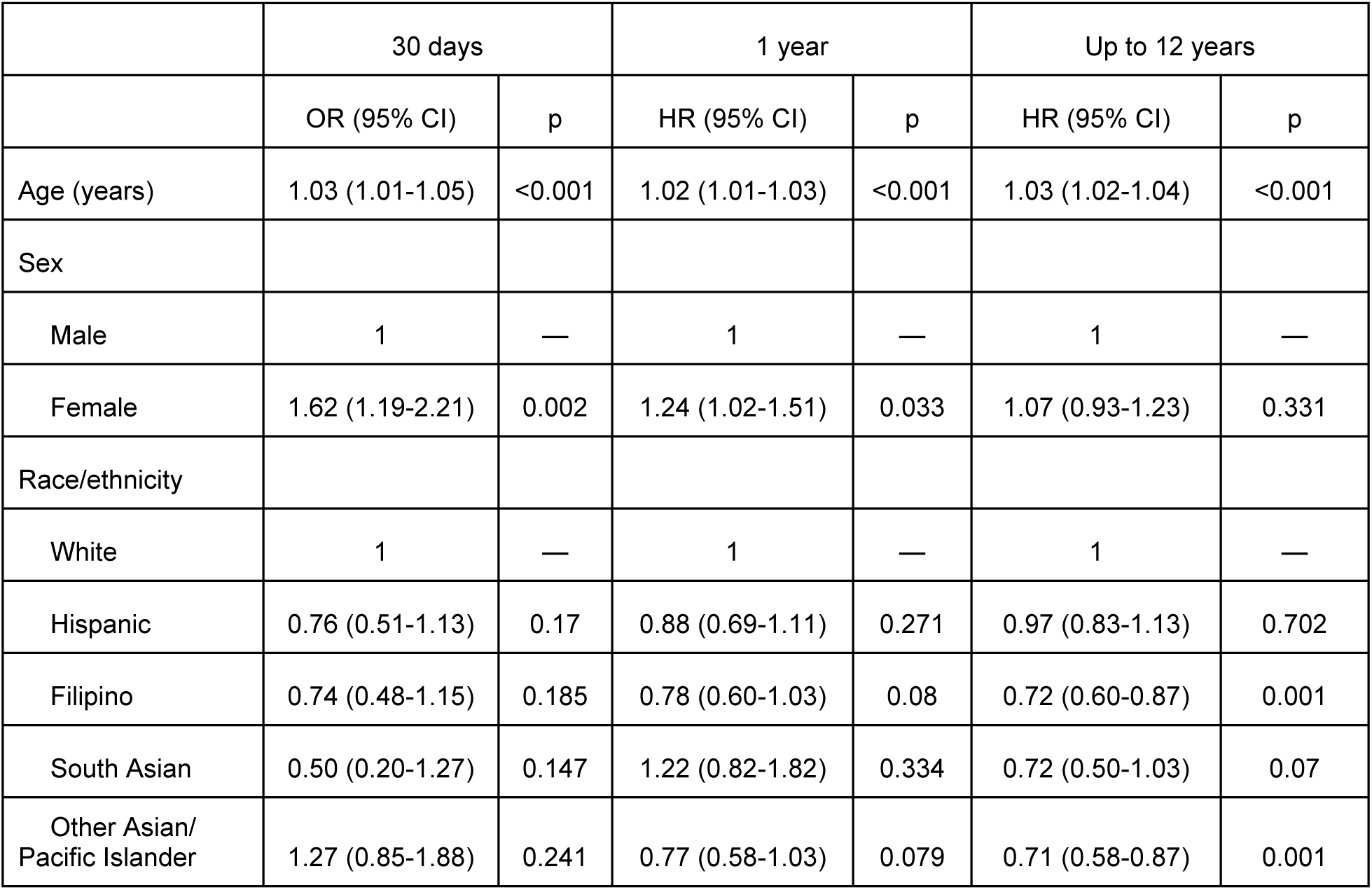
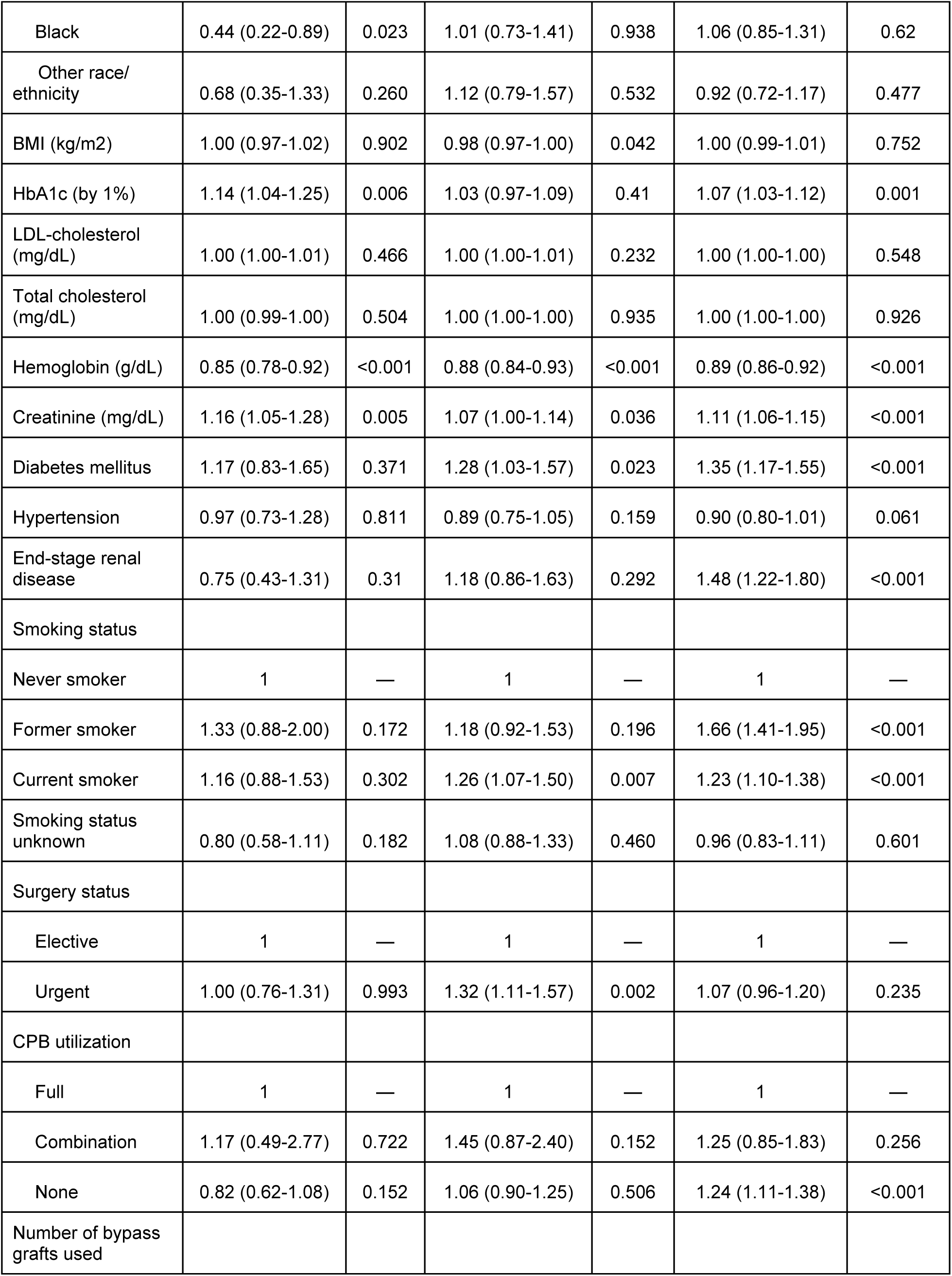
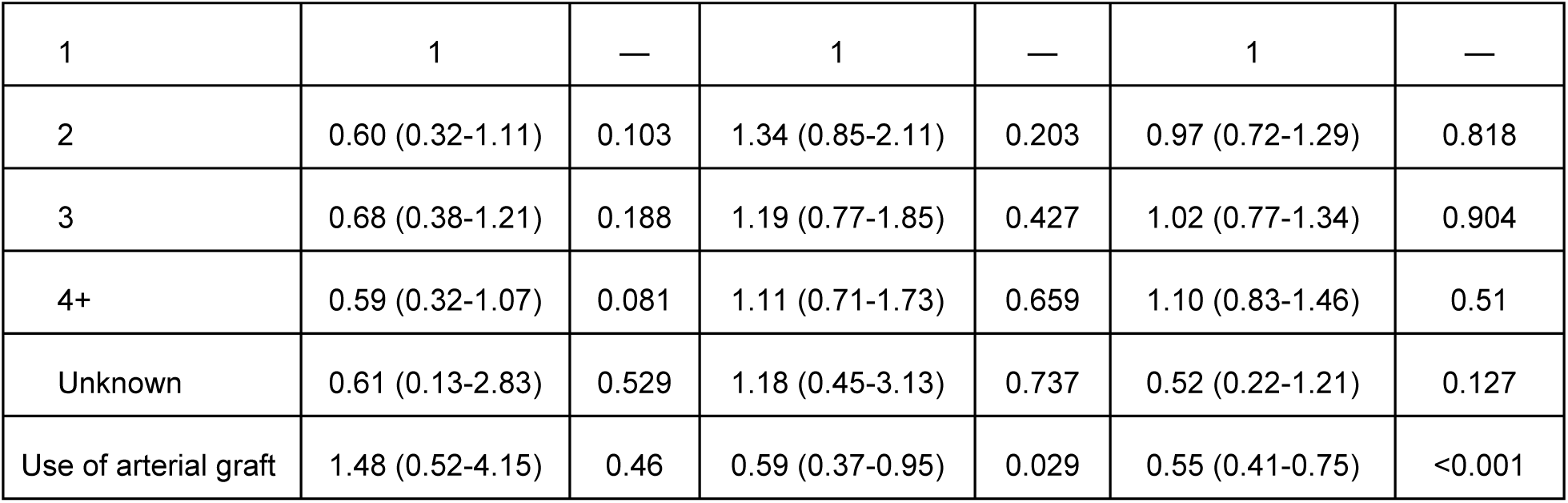
Multivariable analyses in MACE prediction at 30 days, 1 year and 12 years follow up post CABG.

|  | 30 days |  | 1 year |  | Up to 12 years |  |
| --- | --- | --- | --- | --- | --- | --- |
|  | OR (95% CI) | p | HR (95% CI) | p | HR (95% CI) | p |
| Age (years) | 1.03 (1.01-1.05) | <0.001 | 1.02 (1.01-1.03) | <0.001 | 1.03 (1.02-1.04) | <0.001 |
| Sex |  |  |  |  |  |  |
| Male | 1 | — | 1 | — | 1 | — |
| Female | 1.62 (1.19-2.21) | 0.002 | 1.24 (1.02-1.51) | 0.033 | 1.07 (0.93-1.23) | 0.331 |
| Race/ethnicity |  |  |  |  |  |  |
| White | 1 | — | 1 | — | 1 | — |
| Hispanic | 0.76 (0.51-1.13) | 0.17 | 0.88 (0.69-1.11) | 0.271 | 0.97 (0.83-1.13) | 0.702 |
| Filipino | 0.74 (0.48-1.15) | 0.185 | 0.78 (0.60-1.03) | 0.08 | 0.72 (0.60-0.87) | 0.001 |
| South Asian | 0.50 (0.20-1.27) | 0.147 | 1.22 (0.82-1.82) | 0.334 | 0.72 (0.50-1.03) | 0.07 |
| Other Asian/ Pacific Islander | 1.27 (0.85-1.88) | 0.241 | 0.77 (0.58-1.03) | 0.079 | 0.71 (0.58-0.87) | 0.001 |
| Black | 0.44 (0.22-0.89) | 0.023 | 1.01 (0.73-1.41) | 0.938 | 1.06 (0.85-1.31) | 0.62 |
| Other race/<br>ethnicity | 0.68 (0.35-1.33) | 0.260 | 1.12 (0.79-1.57) | 0.532 | 0.92 (0.72-1.17) | 0.477 |
| BMI (kg/m2) | 1.00 (0.97-1.02) | 0.902 | 0.98 (0.97-1.00) | 0.042 | 1.00 (0.99-1.01) | 0.752 |
| HbA1c (by 1%) | 1.14 (1.04-1.25) | 0.006 | 1.03 (0.97-1.09) | 0.41 | 1.07 (1.03-1.12) | 0.001 |
| LDL-cholesterol<br>(mg/dL) | 1.00 (1.00-1.01) | 0.466 | 1.00 (1.00-1.01) | 0.232 | 1.00 (1.00-1.00) | 0.548 |
| Total cholesterol<br>(mg/dL) | 1.00 (0.99-1.00) | 0.504 | 1.00 (1.00-1.00) | 0.935 | 1.00 (1.00-1.00) | 0.926 |
| Hemoglobin (g/dL) | 0.85 (0.78-0.92) | <0.001 | 0.88 (0.84-0.93) | <0.001 | 0.89 (0.86-0.92) | <0.001 |
| Creatinine (mg/dL) | 1.16 (1.05-1.28) | 0.005 | 1.07 (1.00-1.14) | 0.036 | 1.11 (1.06-1.15) | <0.001 |
| Diabetes mellitus | 1.17 (0.83-1.65) | 0.371 | 1.28 (1.03-1.57) | 0.023 | 1.35 (1.17-1.55) | <0.001 |
| Hypertension | 0.97 (0.73-1.28) | 0.811 | 0.89 (0.75-1.05) | 0.159 | 0.90 (0.80-1.01) | 0.061 |
| End-stage renal<br>disease | 0.75 (0.43-1.31) | 0.31 | 1.18 (0.86-1.63) | 0.292 | 1.48 (1.22-1.80) | <0.001 |
| Smoking status |  |  |  |  |  |  |
| Never smoker | 1 | — | 1 | — | 1 | — |
| Former smoker | 1.33 (0.88-2.00) | 0.172 | 1.18 (0.92-1.53) | 0.196 | 1.66 (1.41-1.95) | <0.001 |
| Current smoker | 1.16 (0.88-1.53) | 0.302 | 1.26 (1.07-1.50) | 0.007 | 1.23 (1.10-1.38) | <0.001 |
| Smoking status<br>unknown | 0.80 (0.58-1.11) | 0.182 | 1.08 (0.88-1.33) | 0.460 | 0.96 (0.83-1.11) | 0.601 |
| Surgery status |  |  |  |  |  |  |
| Elective | 1 | — | 1 | — | 1 | — |
| Urgent | 1.00 (0.76-1.31) | 0.993 | 1.32 (1.11-1.57) | 0.002 | 1.07 (0.96-1.20) | 0.235 |
| CPB utilization |  |  |  |  |  |  |
| Full | 1 | — | 1 | — | 1 | — |
| Combination | 1.17 (0.49-2.77) | 0.722 | 1.45 (0.87-2.40) | 0.152 | 1.25 (0.85-1.83) | 0.256 |
| None | 0.82 (0.62-1.08) | 0.152 | 1.06 (0.90-1.25) | 0.506 | 1.24 (1.11-1.38) | <0.001 |
| Number of bypass<br>grafts used |  |  |  |  |  |  |
| 1 | 1 | — | 1 | — | 1 | — |
| 2 | 0.60 (0.32-1.11) | 0.103 | 1.34 (0.85-2.11) | 0.203 | 0.97 (0.72-1.29) | 0.818 |
| 3 | 0.68 (0.38-1.21) | 0.188 | 1.19 (0.77-1.85) | 0.427 | 1.02 (0.77-1.34) | 0.904 |
| 4+ | 0.59 (0.32-1.07) | 0.081 | 1.11 (0.71-1.73) | 0.659 | 1.10 (0.83-1.46) | 0.51 |
| Unknown | 0.61 (0.13-2.83) | 0.529 | 1.18 (0.45-3.13) | 0.737 | 0.52 (0.22-1.21) | 0.127 |
| Use of arterial graft | 1.48 (0.52-4.15) | 0.46 | 0.59 (0.37-0.95) | 0.029 | 0.55 (0.41-0.75) | <0.001 |

At long term follow-up (up to 12 years), increased risk of MACE was significantly associated with age (HR 1.03 [95% CI 1.02-1.04]), increased creatinine (HR 1.11 [95% CI 1.06-1.15]) and increased HbA1c (HR 1.07 [95% CI 1.03-1.12]). Comorbid conditions such as DM (HR 1.35 [95% CI 1.17-1.55]), ESRD (HR 1.48 [95% CI 1.22-1.80]), and former (HR 1.66 [95% CI 1.41-1.95]) or current smoking status (HR 1.23 [95% CI 1.10-1.38]) were also associated with higher MACE at long-term follow-up (Figure 3). Variables associated with decreased incidence of MACE at end of follow-up included Filipino (HR 0.72 [95% CI 0.60-0.87]) or Other Asian/ Pacific Islander (HR 0.71 [95% CI 0.58-0.87]) race/ ethnicity as compared to White race/ethnicity, and increased hemoglobin (HR 0.89 [95% CI 0.86-0.92]). Lack of CPB utilization was associated with higher MACE (HR 1.24 [95% CI 1.11-1.38]) while use of arterial graft was associated with lower MACE (HR 0.55 [95% CI 0.41-0.75]).

**Figure 3.**
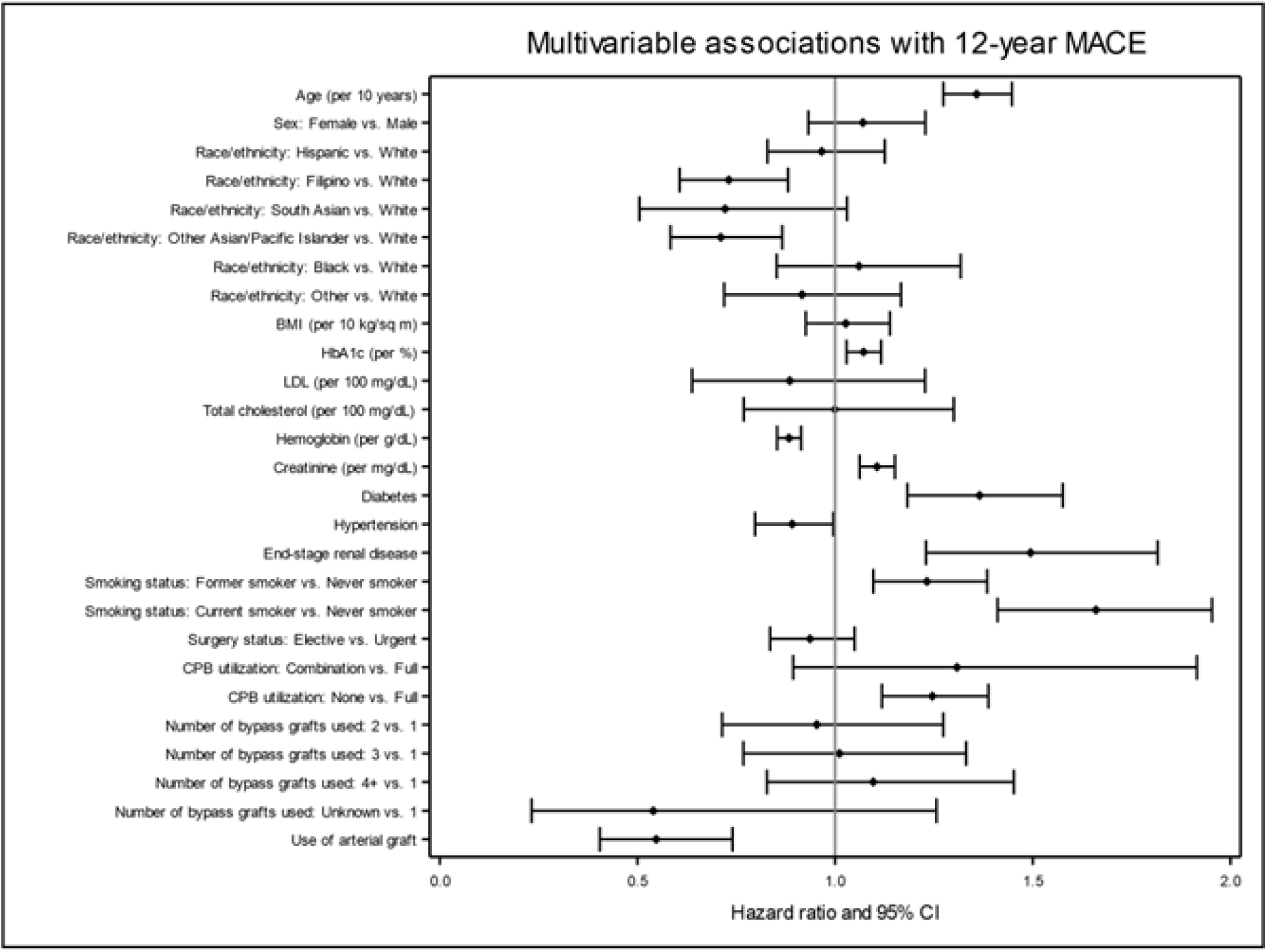
Multivariable associations with 12-year MACE. Forrest plot shows the HR (95% CI) for risk factors in prediction of MACE at 12 years post CABG in this Kaiser Permanente Northern California cohort. Model was adjusted for demographic, clinical and surgical characteristics.

In Kaplan-Meier survival analyses, differences in MACE-free survival at up to 12 years follow-up were observed when stratified by sex (Figure 4, log-rank p <0.001) and by race/ ethnicity (Figure 5, log-rank p<0.001). Lowest MACE-free survival was observed with female sex when compared to male sex (p<0.001), and with Black and Other race/ ethnic group categories, when compared to White race/ethnicity (p<0.001). Higher MACE-free survival was associated with Other Asian/ Pacific Islander, South Asian, and Filipino in comparison to White patients (p<0.001).

**Figure 4.**
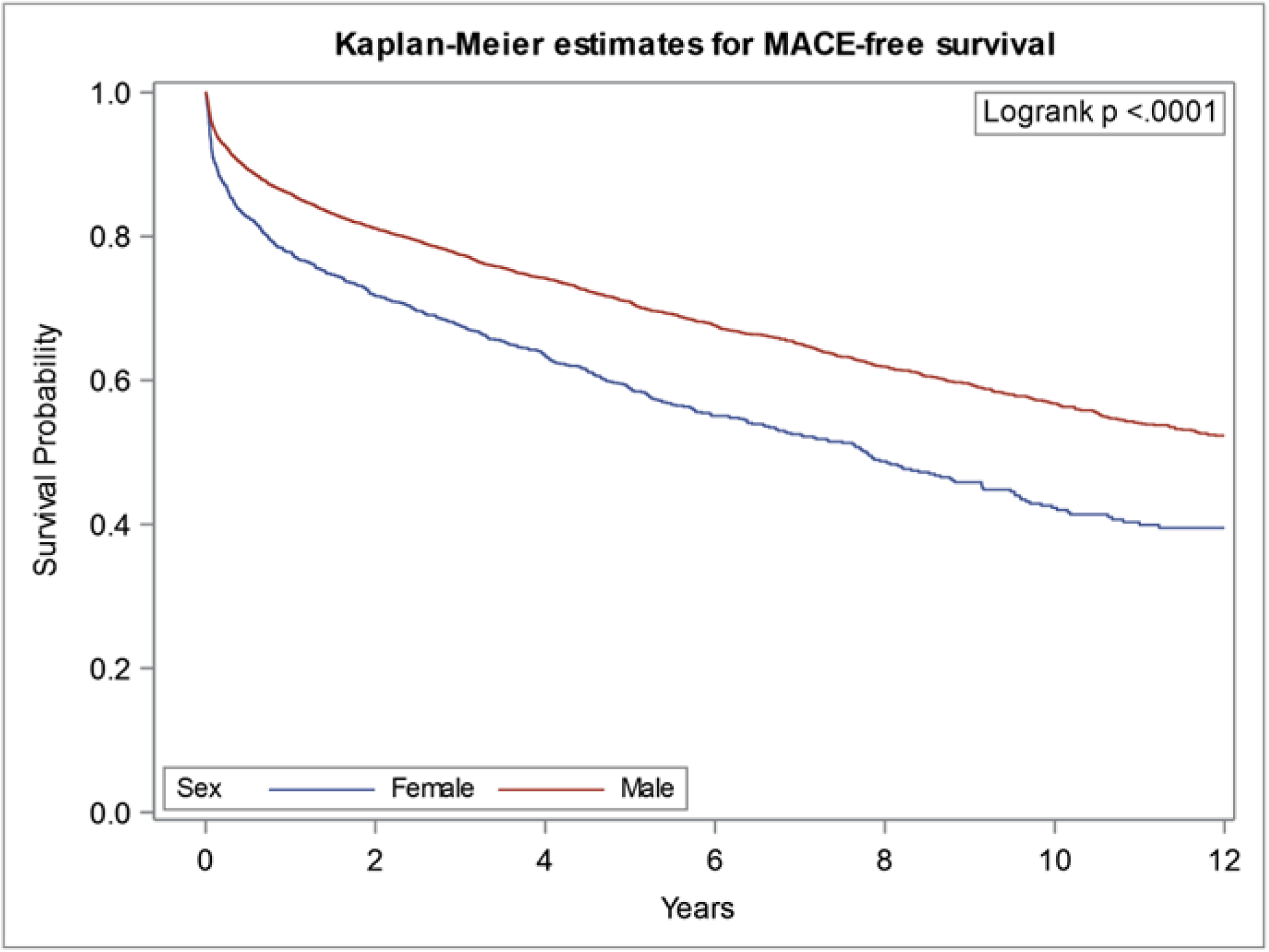
Kaplan-Meier estimates for MACE-free survival following CABG, stratified by sex. MACE was defined as death, MI, stroke, PCI or redo CABG at up to 12 years follow up.

**Figure 5.**
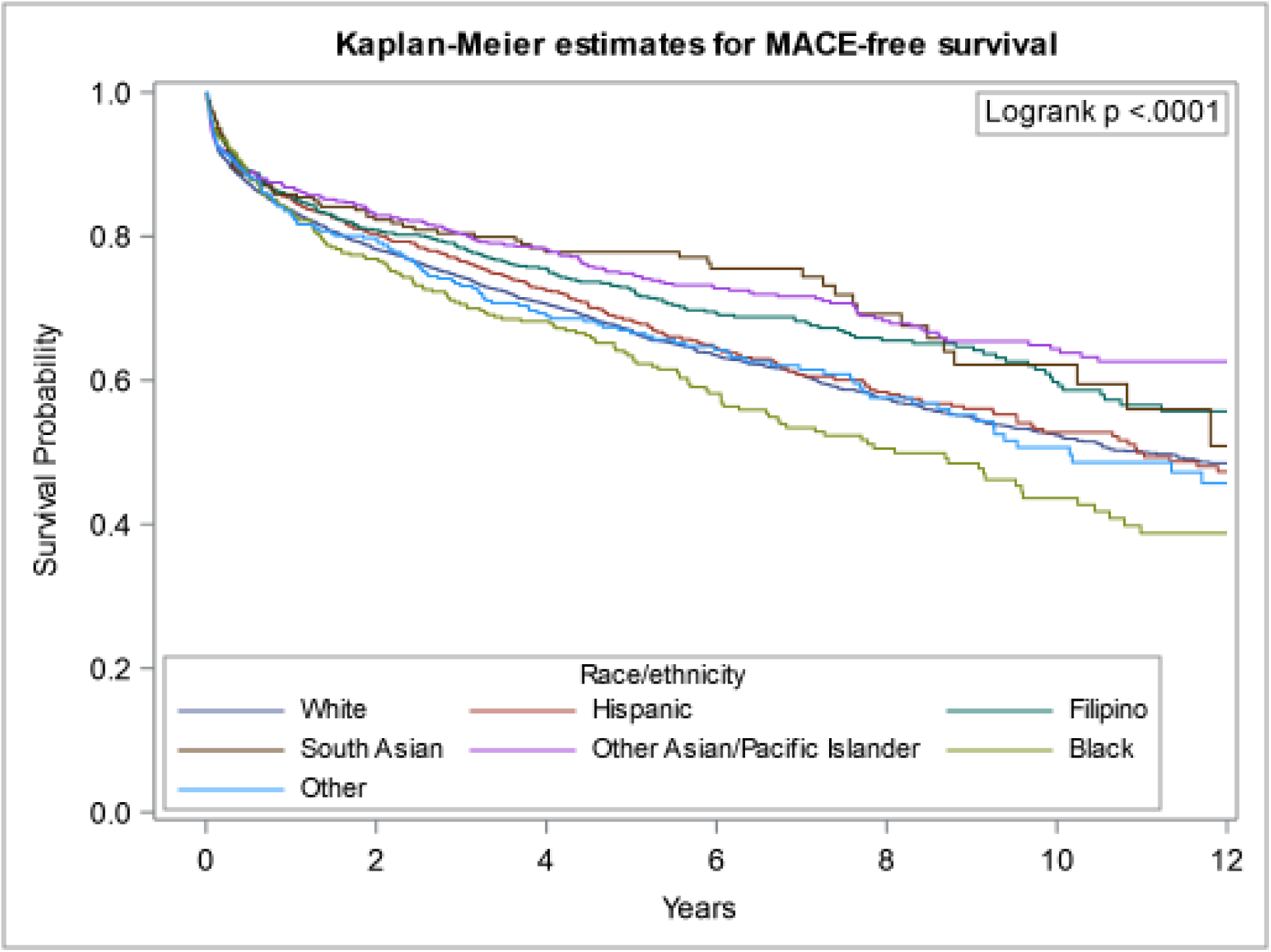
Kaplan-Meier estimates for MACE-free survival following CABG, stratified by race/ ethnicity. MACE was defined as death, MI, stroke, PCI or redo CABG at up to 12 years follow up.

## Discussion

In this large retrospective cohort study following a diverse set of 7405 patients up to 12 years post CABG in an integrated healthcare system, we observed lower MACE free survival in female and Black patients. In a multivariable regression analysis, factors associated with higher hazard of MACE included increasing medical complexity, such as older age, diagnosis of ESRD and DM, as well as surgical characteristics such as lack of CPB use or arterial grafting. These findings corroborate prior studies with known associations with MACE and comorbid cardiac conditions such as DM and ESRD.^30–32^ Of note, sex and race/ ethnicity were not independently associated with higher MACE in multivariable regression analysis, suggesting the burden of intermediary factors including comorbid medical and surgical factors, and social determinants of health to be potential explanations for the observed disparities.

Our findings regarding sex differences in post CABG outcomes remain consistent with existing literature, with female patients experiencing disproportionately higher MACE than male patients.^6,33,34^ This difference is largely driven by preoperative or operative characteristics such as older age with a higher cardiac risk factor burden, and surgical characteristics leading to lower use of arterial grafts, higher rate of graft failure and lower likelihood of complete revascularization.^6,35^ In our study, females were older with greater frequency of prior MI, DM, HTN, ESRD and higher STS score and more likely to be urgent, compared to elective, surgical status. Available census tract data also suggests differences in social determinants of health, with females less likely to be college graduates and have lower median household incomes, though limited by individual-level data.

Similar to our study findings, Black individuals have historically demonstrated worse outcomes post CABG, which can be partially explained by the presence of higher cardiac comorbidities. In the NHANES survey data, Black individuals have higher rates of DM and exhibit a higher hazard ratio of developing ERD relative to White individuals even after adjusting for GFR.^36^ Black individuals also had higher rates of HTN and heart failure, leading to higher disease burden at a younger age.^25,37^ Studies have shown correlations between racial discrimination with higher coronary artery calcium score, high-sensitive C-reactive protein and HTN akin to chronic systemic inflammation and stress.^36,38^ In our study cohort, Black patients represented the highest female population (34% versus 18% in total cohort) and had the greatest frequency of current and former smokers, highest BMI, LDL-c and creatinine values, and the lowest hemoglobin values. Black patients were among the higher STS scores and more likely to have an urgent, versus elective, surgical status, which has been similarly noted in other studies.^4,13,36^ Understanding the upstream mechanisms for these racial disparities requires greater emphasis on social and structural factors that contribute to inequitable outcomes.

Differences in social determinants such as education level, healthcare literacy, English proficiency, and financial stability, can lead to differences in prevalence, diagnosis and management of comorbid conditions as well as access to timely and appropriate care. Black patients are less likely to undergo CABG with lower referral rates than White patients.^4,13,36^ In a national retrospective cohort study evaluating 1429 acute hospitals, Black individuals consistently had lower access to CABG regardless of healthcare system or hospital, which may be due to implicit biases rooted in race, socioeconomic and/ or education differences.^9^ Furthermore, Black and Hispanic patients consistently receive less risk reduction counseling or referral to cardiac rehabilitation post CABG compared to White patients, with demonstrated poor long-term survival rates disproportionately affecting younger Black patients.^36^ Finally, while current clinical prediction algorithms include race/ ethnicity and sex as independent variables to predict post cardiovascular surgery morbidity and mortality, they do not include granular race/ ethnicity and social determinants of health, which are powerful prognosticators of postoperative outcomes.^39,40^ Likewise, global gender inequalities still exist, with female patients less likely to have food and healthcare security, higher education, and financial independence.^36,41^ Female patients often carry a significant burden in terms of childcare and child rearing, and are more vulnerable to competing priorities in relation to their personal medical care and participation in research trials, potentially leading to under-diagnosis of relevant cardiac conditions in a timely manner.^36,41^ Understanding upstream contributors to this lack of timely diagnosis of cardiac conditions in females requires a multi-faceted approach evaluating health perceptions and disparities in care delivery.

There are several limitations to this study. As this study is retrospective, data extraction was limited to electronic health record review, and comprehensive, individual-level social determinants of health data was unavailable. Race/ ethnicity was based on self-identification, and biologically assigned sex at birth was extracted from the medical record (gender identity was not studied). The Kaiser Permanente Northern California cohort, while representative of the racial/ ethnic and socioeconomic diversity in the state, may have overrepresentation of Asians and Other Asians/ Pacific Islanders, and underrepresentation of White, Black, and Hispanic patients when compared to statistics from the United States Census Bureau.^42^ Furthermore, grouping of Native Americans, multi-ethnic groups, and missing race/ ethnicity data into an “Other” group may have prevented adequate detection of potential disparities in this heterogeneous population, though data in these groups is limited by small numbers. Strengths of this study include the large cohort size, long-term follow up based on membership, and use of a diverse integrated healthcare system, where variability in access to care would be theoretically limited. We also provide granular race/ ethnicity data, particularly with disaggregation of a heterogeneous Asian population, to help further understand outcome disparities.

In this large retrospective cohort study from an integrated healthcare system, disparities in post CABG outcomes persist by sex and race/ ethnicity. Focused contemporary efforts are required to reduce upstream risk factor burden in high risk groups in order to reduce these postoperative disparities.

## Data Availability

Requests to access the dataset from qualified researchers trained in human subjects? confidentiality protocols may be sent to the principal investigator. The patient data is owned by the Kaiser Foundation Health Plan, Inc., Kaiser Foundation Hospitals, Inc., and The Permanente Medical Group, Inc. Because of their third-party rights, it is not possible to make the data publicly available without restriction.?

## Non-standard Abbreviations and Acronyms

CABG: Coronary artery bypass graft surgery
HDL-c: High density lipoprotein cholesterol
LDL-c: Low density lipoprotein cholesterol
MACE: Major adverse cardiovascular event
OR: Odds ratio
PCI: Percutaneous coronary intervention

## Acknowledgments

All authors have acknowledged and approved their contributions to this manuscript.

## References

1. Mensah GA, Roth GA, Fuster V. The Global Burden of Cardiovascular Diseases and Risk Factors: 2020 and Beyond. J Am Coll Cardiol. 2019;74:2529–2532.

2. Serruys PW, Morice M-C, Kappetein AP, Colombo A, Holmes DR, Mack MJ, Ståhle E, Feldman TE, Brand M van den, Bass EJ, Dyck NV, Leadley K, Dawkins KD, Mohr FW. Percutaneous Coronary Intervention versus Coronary-Artery Bypass Grafting for Severe Coronary Artery Disease. N Engl J Med. 2009;360:961–972.

3. Lawton JS, Tamis-Holland JE, Bangalore S, Bates ER, Beckie TM, Bischoff JM, Bittl JA, Cohen MG, DiMaio JM, Don CW, Fremes SE, Gaudino MF, Goldberger ZD, Grant MC, Jaswal JB, Kurlansky PA, Mehran R, Metkus TS, Nnacheta LC, Rao SV, Sellke FW, Sharma G, Yong CM, Zwischenberger BA. 2021 ACC/AHA/SCAI Guideline for Coronary Artery Revascularization: Executive Summary: A Report of the American College of Cardiology/American Heart Association Joint Committee on Clinical Practice Guidelines. Circulation. 2022;145:e4–e17.

4. Benedetto U, Kamel MK, Khan FM, Angelini GD, Caputo M, Girardi LN, Gaudino M. Are racial differences in hospital mortality after coronary artery bypass graft surgery real? A risk-adjusted meta-analysis. J Thorac Cardiovasc Surg. 2019;157:2216–2225.e4.

5. Bridges CR, Edwards FH, Peterson ED, Coombs LP. The effect of race on coronary bypass operative mortality. J Am Coll Cardiol. 2000;36:1870–1876.

6. Angraal S, Khera R, Wang Y, Lu Y, Jean R, Dreyer RP, Geirsson A, Desai NR, Krumholz HM. Sex and Race Differences in the Utilization and Outcomes of Coronary Artery Bypass Grafting Among Medicare Beneficiaries, 1999–2014. J Am Heart Assoc Cardiovasc Cerebrovasc Dis. 2018;7:e009014.

7. Institute of Medicine (US) Committee on Understanding and Eliminating Racial and Ethnic Disparities in Health Care. Unequal Treatment: Confronting Racial and Ethnic Disparities in Health Care. Washington (DC): National Academies Press (US); 2003. Available at http://www.ncbi.nlm.nih.gov/books/NBK220358/. Accessed July 16, 2024.

8. Milam AJ, Ogunniyi MO, Faloye AO, Castellanos LR, Verdiner RE, Stewart JW, Chukumerije M, Okoh AK, Bradley S, Roswell RO, Douglass PL, Oyetunji SO, Iribarne A, Furr-Holden D, Ramakrishna H, Hayes SN. Racial and Ethnic Disparities in Perioperative Health Care Among Patients Undergoing Cardiac Surgery: JACC State-of-the-Art Review. J Am Coll Cardiol. 2024;83:530–545.

9. Ayanian JZ, Udvarhelyi IS, Gatsonis CA, Pashos CL, Epstein AM. Racial differences in the use of revascularization procedures after coronary angiography. JAMA. 1993;269:2642–2646.

10. Hannan EL, van Ryn M, Burke J, Stone D, Kumar D, Arani D, Pierce W, Rafii S, Sanborn TA, Sharma S, Slater J, DeBuono BA. Access to coronary artery bypass surgery by race/ethnicity and gender among patients who are appropriate for surgery. Med Care. 1999;37:68–77.

11. Johnson PA, Lee TH, Cook EF, Rouan GW, Goldman L. Effect of race on the presentation and management of patients with acute chest pain. Ann Intern Med. 1993;118:593–601.

12. Petersen LA, Wright SM, Peterson ED, Daley J. Impact of race on cardiac care and outcomes in veterans with acute myocardial infarction. Med Care. 2002;40:I86–96.

13. Zea-Vera R, Asokan S, Shah RM, Ryan CT, Chatterjee S, Wall MJ, Coselli JS, Rosengart TK, Kayani WT, Jneid H, Ghanta RK. Racial/ethnic differences persist in treatment choice and outcomes in isolated intervention for coronary artery disease. J Thorac Cardiovasc Surg. 2023;166:1087–1096.e5.

14. Rangrass G, Ghaferi AA, Dimick JB. Explaining racial disparities in outcomes after cardiac surgery: the role of hospital quality. JAMA Surg. 2014;149:223–227.

15. Tabata M, Grab JD, Khalpey Z, Edwards FH, O’Brien SM, Cohn LH, Bolman RM. Prevalence and variability of internal mammary artery graft use in contemporary multivessel coronary artery bypass graft surgery: analysis of the Society of Thoracic Surgeons National Cardiac Database. Circulation. 2009;120:935–940.

16. O’Shaughnessy S, Tangel V, Dzotsi S, Jiang S, White R, Hoyler M. Non-White Race/Ethnicity and Female Sex Are Associated with Increased Allogeneic Red Blood Cell Transfusion in Cardiac Surgery Patients: 2007-2018. J Cardiothorac Vasc Anesth. 2022;36:1908–1918.

17. Padmanabhan H, Brookes MJ, Nevill AM, Luckraz H. Association Between Anemia and Blood Transfusion With Long-term Mortality After Cardiac Surgery. Ann Thorac Surg. 2019;108:687–692.

18. Jannati M, Navaei MR, Ronizi LG. A comparative review of the outcomes of using arterial versus venous conduits in coronary artery bypass graft (CABG). J Fam Med Prim Care. 2019;8:2768–2773.

19. Salmasi MY, Ravishankar R, Abdullahi Y, Hartley P, Kyriazis PG, Athanasiou T, Punjabi P. Predictors of outcome after CABG in the South-Asian community: a propensity matched analysis. Perfusion. 2023;38:75–84.

20. Volgman A, Palaniappan L, Aggarwal N, Gupta M, Khandelwal A, Krishnan A, Lichtman J, Mehta L, Patel H, Shah K, Shah S, AHA Council on Epidemiology and Prevention, Cardiovascular Disease and Stroke in Women and Special Populations Committee of the Council on Clinical Cardiology, Council on Cardiovascular and Stroke Nursing, Council on Quality of Care and Outcomes Research, Stroke Council. Atherosclerotic Cardiovascular Disease in South Asians in the United States: Epidemiology, Risk Factors, and Treatments: A Scientific Statement From the American Heart Association. Circulation. 2018;3:e1–e34.

21. Brister SJ, Hamdulay Z, Verma S, Maganti M, Buchanan MR. Ethnic diversity: South Asian ethnicity is associated with increased coronary artery bypass grafting mortality. J Thorac Cardiovasc Surg. 2007;133:150–154.

22. Becker ER, Rahimi A. Disparities in race/ethnicity and gender in in-hospital mortality rates for coronary artery bypass surgery patients. J Natl Med Assoc. 2006;98:1729–1739.

23. Deb S, Wijeysundera HC, Ko DT, Tsubota H, Hill S, Fremes SE. Coronary artery bypass graft surgery vs percutaneous interventions in coronary revascularization: a systematic review. JAMA. 2013;310:2086–2095.

24. Kilic A, Higgins RSD, Whitson BA, Kilic A. Racial disparities in outcomes of adult heart transplantation. Circulation. 2015;131:882–889.

25. Enumah ZO, Canner JK, Alejo D, Warren DS, Zhou X, Yenokyan G, Matthew T, Lawton JS, Higgins RSD. Persistent Racial and Sex Disparities in Outcomes After Coronary Artery Bypass Surgery. Ann Surg. 2020;272:660–667.

26. Benjamin EJ, Muntner P, Alonso A, Bittencourt MS, Callaway CW, Carson AP, Chamberlain AM, Chang AR, Cheng S, Das SR, Delling FN, Djousse L, Elkind MSV, Ferguson JF, Fornage M, Jordan LC, Khan SS, Kissela BM, Knutson KL, Kwan TW, Lackland DT, Lewis TT, Lichtman JH, Longenecker CT, Loop MS, Lutsey PL, Martin SS, Matsushita K, Moran AE, Mussolino ME, O’Flaherty M, Pandey A, Perak AM, Rosamond WD, Roth GA, Sampson UKA, Satou GM, Schroeder EB, Shah SH, Spartano NL, Stokes A, Tirschwell DL, Tsao CW, Turakhia MP, VanWagner LB, Wilkins JT, Wong SS, Virani SS, On behalf of the American Heart Association Council on Epidemiology and Prevention Statistics Committee and Stroke Statistics Subcommittee. Heart Disease and Stroke Statistics—2019 Update: A Report From the American Heart Association. Circulation. 2019;139. doi:10.1161/CIR.0000000000000659.

27. Gaudino M, Chadow D, Rahouma M, Soletti GJ, Sandner S, Perezgrovas-Olaria R, Audisio K, Cancelli G, Bratton BA, Fremes S, Kurlansky P, Girardi L, Habib RH. Operative Outcomes of Women Undergoing Coronary Artery Bypass Surgery in the US, 2011 to 2020. JAMA Surg. 2023;158:494–502.

28. Alam M, Bandeali SJ, Kayani WT, Ahmad W, Shahzad SA, Jneid H, Birnbaum Y, Kleiman NS, Coselli JS, Ballantyne CM, Lakkis N, Virani SS. Comparison by meta-analysis of mortality after isolated coronary artery bypass grafting in women versus men. Am J Cardiol. 2013;112:309–317.

29. Hassan A, Chiasson M, Buth K, Hirsch G. Women have worse long-term outcomes after coronary artery bypass grafting than men. Can J Cardiol. 2005;21:757–762.

30. Endo D, Yamamoto T, Kajimoto K, Matsushita S, Dohi S, Shimada A, Yokoyama Y, Io H, Suzuki Y, Tabata M, Amano A. Coronary Artery Bypass Grafting in Patients with Chronic Kidney Disease: Chronic Kidney Disease Has an Independent Adverse Effect on the Long-Term Outcome of Coronary Artery Bypass Grafting. BioMed Res Int. 2022;2022:4994970.

31. Kusu-Orkar T-E, Kermali M, Oguamanam N, Bithas C, Harky A. Coronary artery bypass grafting: Factors affecting outcomes. J Card Surg. 2020;35:3503–3511.

32. Zafrir B, Leviner DB, Saliba W, Sharoni E. Prognostic Interplay of Chronic Kidney Disease, Anemia, and Diabetes in Coronary Bypass Surgery. Ann Thorac Surg. 2021;111:94–101.

33. Gaudino M, Di Franco A, Alexander JH, Bakaeen F, Egorova N, Kurlansky P, Boening A, Chikwe J, Demetres M, Devereaux PJ, Diegeler A, Dimagli A, Flather M, Hameed I, Lamy A, Lawton JS, Reents W, Robinson NB, Audisio K, Rahouma M, Serruys PW, Hara H, Taggart DP, Girardi LN, Fremes SE, Benedetto U. Sex differences in outcomes after coronary artery bypass grafting: a pooled analysis of individual patient data. Eur Heart J. 2021;43:18–28.

34. Bryce Robinson N, Naik A, Rahouma M, Morsi M, Wright D, Hameed I, Di Franco A, Girardi LN, Gaudino M. Sex differences in outcomes following coronary artery bypass grafting: a meta-analysis. Interact Cardiovasc Thorac Surg. 2021;33:841–847.

35. Harik L, Perezgrovas-Olaria R, Jr Soletti G, Dimagli A, Alzghari T, An KR, Cancelli G, Gaudino M. Sex differences in coronary artery bypass graft surgery outcomes: a narrative review. J Thorac Dis. 2023;15:5041–5054.

36. Morris AA, Masoudi FA, Abdullah AR, Banerjee A, Brewer LC, Commodore-Mensah Y, Cram P, DeSilvey SC, Hines AL, Ibrahim NE, Jackson EA, Joynt Maddox KE, Makaryus AN, Piña IL, Rodriguez-Monserrate CP, Roger VL, Thorpe FF, Williams KA. 2024 ACC/AHA Key Data Elements and Definitions for Social Determinants of Health in Cardiology: A Report of the American College of Cardiology/American Heart Association Joint Committee on Clinical Data Standards. Circ Cardiovasc Qual Outcomes. 2024;:e000133.

37. NHANES - Interactive Data Visualizations. Available at https://www.cdc.gov/nchs/nhanes/visualization/index.htm. Accessed July 16, 2024.

38. Sims KD, Sims M, Glover LM, Smit E, Odden MC. Perceived Discrimination and Trajectories of C-Reactive Protein: The Jackson Heart Study. Am J Prev Med. 2020;58:199–207.

39. Shawon MSR, Odutola M, Falster MO, Jorm LR. Patient and hospital factors associated with 30-day readmissions after coronary artery bypass graft (CABG) surgery: a systematic review and meta-analysis. J Cardiothorac Surg. 2021;16:172.

40. Anderson JE, Li Z, Romano PS, Parker J, Chang DC. Should Risk Adjustment for Surgical Outcomes Reporting Include Sociodemographic Status? A Study of Coronary Artery Bypass Grafting in California. J Am Coll Surg. 2016;223:221–230.

41. Balafa O, Fernandez-Fernandez B, Ortiz A, Dounousi E, Ekart R, Ferro CJ, Mark PB, Valdivielso JM, Del Vecchio L, Mallamaci F. Sex disparities in mortality and cardiovascular outcomes in chronic kidney disease. Clin Kidney J. 2024;17:sfae044.

42. Sujata null, Thakur R. Unequal burden of equal risk factors of diabetes between different gender in India: a cross-sectional analysis. Sci Rep. 2021;11:22653.

